# Tracing the Benign Nature of Pilocytic Astrocytoma with a Focus on How Gliogenic Regulators Impact its Tumorigenesis: a systematic review

**DOI:** 10.1101/2025.05.09.25327290

**Authors:** Ovais Shafi, Raveena

## Abstract

**Objective:** The objective of this study is to determine how gliogenic regulators impact the Pilocytic Astrocytoma tumorigenesis and what it has to do with the benign nature of Pilocytic Astrocytoma.

**Background:** Pilocytic Astrocytoma (PA) is a benign pediatric brain tumor driven by gliogenic signaling dysregulation, particularly the MAPK/ERK pathway. Unlike high-grade gliomas, PA maintains differentiation cues that limit malignancy. Investigating gliogenic transcription factors, tumor suppressors, and signaling pathways can reveal how PA sustains proliferation without aggressive transformation. This research bridges oncogenic drivers and gliogenic regulation, offering insights into potential therapies and strategies to prevent malignant progression in gliomas.

**Methods:** Databases, including PubMed, MEDLINE, Google Scholar, and open access/ subscription-based journals were searched for articles without any date restrictions, to investigate how gliogenic regulators impact the key PA tumorigenesis regulators and what it has to do with the benign nature of Pilocytic Astrocytoma. Based on the criteria mentioned in the methods section, studies were systematically reviewed to investigate the research question. This study adheres to relevant PRISMA guidelines (Preferred Reporting Items for Systematic Reviews and Meta-Analyses).

**Results:** This study reveals that Pilocytic Astrocytoma (PA) originates through gliogenic differentiation-linked oncogenesis, primarily driven by MAPK/ERK pathway hyperactivation via BRAF fusion or NF1 loss. Despite oncogenic signaling, PA remains benign due to intact PTEN, TP53, and differentiation-linked transcription factors (SOX10, NFIs, PAX6), which suppress uncontrolled proliferation. The tumor microenvironment further restricts angiogenesis and invasion, reinforcing its low malignancy risk. Key glial regulatory pathways, including Notch, Shh, and JAK/STAT, contribute to sustained astrocytic differentiation rather than dedifferentiation. Additionally, PI3K/AKT/mTOR signaling remains tightly controlled, preventing high-grade transformation.

These findings refine our understanding of benign glioma oncogenesis, distinguishing PA from aggressive gliomas like glioblastoma. Identifying molecular players in PA’s stability offers potential biomarkers for diagnosis and suggests differentiation-inducing therapies as a promising approach to limiting tumor progression.

**Conclusion:** Pilocytic Astrocytoma (PA) remains benign due to controlled MAPK/ERK activation, gliogenic transcription factors (PAX6, NFIA, STAT3, SOX10) promoting differentiation, and limited activation of oncogenic pathways (PI3K/AKT, JAK/STAT, Notch, Shh). Low invasiveness, restricted angiogenesis, and a non-immunosuppressive microenvironment prevent malignant transformation. PA lacks genetic instability, avoiding mutations that drive high-grade gliomas. Its gliogenic framework ensures regulated growth, differentiation, and genomic stability, preventing dedifferentiation or invasiveness, solidifying PA as a benign, non-malignant glioma with stable cellular behavior and limited tumor progression potential.

## Background

Pilocytic Astrocytoma (PA) is the most common pediatric brain tumor, characterized by a slow-growing, well-circumscribed nature and a predominantly benign clinical course [1]. Unlike high-grade gliomas, PA rarely undergoes malignant transformation, yet the mechanisms underlying its oncogenesis and restrained tumor progression remain poorly understood. A key aspect of PA biology is its strong association with dysregulated gliogenic signaling, particularly involving the MAPK/ERK pathway, due to alterations such as BRAF fusion or NF1 loss. However, the interaction between oncogenic signaling and the gliogenic framework governing glial cell proliferation and differentiation remains inadequately explored [2].

Understanding how gliogenic transcription factors (e.g., PAX6, NFIA, SOX10), signaling pathways (JAK/STAT, PI3K/AKT/mTOR, Notch, BMP), and tumor suppressors (PTEN, TP53) influence PA development is critical [3]. These players regulate glial progenitor maintenance, astrocytic differentiation, and cell cycle exit, processes that remain intact enough in PA to prevent aggressive transformation. Investigating their roles in PA oncogenesis can elucidate why PA sustains a proliferative but non-malignant state, distinguishing it from high-grade gliomas [4].

This study is essential for bridging the gap between oncogenic drivers and gliogenic regulation in PA. By analyzing how gliogenic mechanisms partially suppress dedifferentiation and invasion, this research offers insights into potential therapeutic strategies that target proliferative signals without disrupting differentiation cues. Moreover, understanding PA’s benign nature could reveal new strategies for preventing malignant progression in other gliomas, making this research highly relevant for neuro-oncology [5].

## Methods

### Aim of the Study

This study investigates how genes, transcription factors, and signaling pathways (BRAF, NF1, FGFR1, PTEN, TP53, SOX10, NFI family (NFIA, NFIB, NFIX), STAT3, PAX6, AP-1, Olig2, MAPK/ERK, PI3K/AKT/mTOR, JAK/STAT, Shh, and Notch) involved in Pilocytic Astrocytoma (PA) development are impacted by gliogenic regulators. The gliogenic regulators examined include IL-6 family, FGFR3, JAK-STAT signaling, BMPs, Notch signaling, Notch effector protein NFIA, SOX9, SOX4, STAT3, GFAP and S100, BMP with SMAD, p300/CBP, Notch/Hey1, HES genes, STAT3-JAK2 interactions, SHH, PAX6 with Nkx6.1, NF-κB signaling, Neuregulin-1, neuronal restrictive silencing factor (NRSF/REST), MAPK, MEK, TGF-β, E2F, TCFL2, p130 with JAK/STAT, transcription factors NFIC and HOPX, Ephrins (EFNB1), and Netrins. These are also the limitations of the study.

The study aims to determine:

- The mechanisms that contribute to the benign nature of Pilocytic Astrocytoma
- Mechanisms by which gliogenic regulators influence PA tumorigenesis via transcription factors and signaling pathways.
- The interaction between gliogenic regulators and tumorigenesis regulators in PA.

### Research Question

How do gliogenic regulators (IL-6 family, FGFR3, JAK-STAT signaling, BMPs, Notch, NFIA, SOX9, SOX4, STAT3, and others) impact the key tumorigenesis factors involved in Pilocytic Astrocytoma (PA) development, and how it regulates the benign nature of Pilocytic Astrocytoma?

### Search Focus

A comprehensive literature search was conducted using the PUBMED database, MEDLINE database, and Google Scholar, as well as open access and subscription-based journals. There were no date restrictions for published articles. The search strategy targeted:

- Key genetic alterations and transcription factor involvement in PA development.
- Interactions between gliogenic regulators and tumor landscape of PA
- Effects of signaling pathways (MAPK/ERK, PI3K/AKT/mTOR, JAK/STAT, Shh, Notch) on PA progression.
- The influence of tumor microenvironment factors on PA proliferation and differentiation.

Screening of the literature was also done on this same basis and related data was extracted. Literature search began in December 2020 and ended in December 2024. An in-depth investigation was conducted during this duration based on the parameters of the study as defined above. During revision, further literature was searched and referenced until March 2025. The literature search and all sections of the manuscript were checked multiple times during the months of revision (January 2025 – March 2025) to maintain the highest accuracy possible. This comprehensive approach ensured that the selected studies provided valuable insights into the gliogenic regulators and their impact on key PA tumorigenesis regulators. This study adheres to relevant PRISMA guidelines (Preferred Reporting Items for Systematic Reviews and Meta-Analyses).

### Search Queries/Keywords

1. Genes and Transcription Factors in PA Development:

- “BRAF mutations in Pilocytic Astrocytoma”
- “NF1 and PA tumorigenesis”
- “FGFR1 signaling in PA”
- “PTEN loss in PA”
- “TP53 and gliogenesis”
- “SOX10 and PA differentiation”
- “NFI family (NFIA, NFIB, NFIX) in gliogenesis”
- “STAT3 activation in PA”
- “PAX6 and PA development”
- “AP-1 in astrocytic tumors”
- “Olig2 and glioma cell fate”
2. Signaling Pathways and PA Progression:

- “MAPK/ERK pathway in PA”
- “PI3K/AKT/mTOR signaling in PA”
- “JAK/STAT pathway in astrocytoma”
- “Shh signaling in glioma”
- “Notch signaling and PA growth”
3. Gliogenic Regulators and PA Tumorigenesis:

- “IL-6 family cytokines in gliogenesis”
- “FGFR3 mutations and PA differentiation”
- “BMP-SMAD signaling in PA”
- “Notch effector protein NFIA and PA”
- “SOX9, SOX4 in PA stemness”
- “STAT3-JAK2 interactions in astrocytoma”
- “GFAP and S100 expression in PA”
- “TGF-β and gliogenesis in PA”
- “Ephrins (EFNB1) and Netrins in PA progression”

Boolean operators (AND, OR) were used to refine search results. Additional searches were conducted to identify studies on gliogenic regulators and its effects on PA tumor progression.

### Objectives of the Search

1. To investigate how gliogenic regulators influence the expression and function of key genes, transcription factors and signaling pathways in PA.
2. To assess the benign nature of PA tumorigenesis.
3. To determine how these dysregulations, contribute to PA progression and stemness alterations.

### Screening and Eligibility Criteria

Initial Screening: Titles and abstracts were reviewed to ensure relevance to PA tumorigenesis, gliogenic regulators, and benign nature of PA.

Full-Text Review: Articles were further examined for detailed mechanistic insights into the involvement of genes, transcription factors and signaling pathways in PA development.

Data Extraction:

Studies were categorized based on their focus on:

- Mechanisms underlying PA development.
- The impact of gliogenic regulators on PA cell fate.
- Benign nature of PA tumorigenesis.

### Inclusion and Exclusion Criteria

Inclusion Criteria:

- Studies analyzing the role of BRAF, NF1, FGFR1, PTEN, TP53, SOX10, and other transcription factors in PA.
- Research exploring the impact of gliogenic regulators (IL-6 family, BMPs, Notch, NFIA, STAT3, etc.) on PA development.
- Studies detailing interactions between gliogenic regulators, inflammation, and PA progression.
- Studies involving astrocytic tumors with specific reference to gliogenesis-associated transcriptional regulation.

Exclusion Criteria:

- Articles that did not conform to the study focus.
- Insufficient methodological rigor.
- Data not aligning with the research questions.

### Rationale for Screening and Inclusion

- BRAF, NF1, FGFR1, PTEN, TP53: Well-established genetic drivers in PA.
- SOX10, NFIA, NFIB, NFIX: Key regulators of gliogenic differentiation in PA.
- STAT3, PAX6, AP-1, Olig2: Transcription factors essential for PA progression and astrocytic fate determination.
- MAPK/ERK, PI3K/AKT/mTOR, JAK/STAT, Shh, Notch: Crucial signaling pathways implicated in PA proliferation and survival.
- Gliogenic regulators (IL-6, BMPs, Notch, STAT3, NFIA, SOX9, etc.): Modulate transcriptional networks driving gliogenesis in PA.

This comprehensive screening and inclusion rationale ensure that the selected studies provide valuable insights into the roles of these critical genes, transcription factors, and signaling pathways to understand the benign nature of Pilocytic Astrocytoma.

PRISMA Flow Diagram is Fig 1.

**Fig 1.**
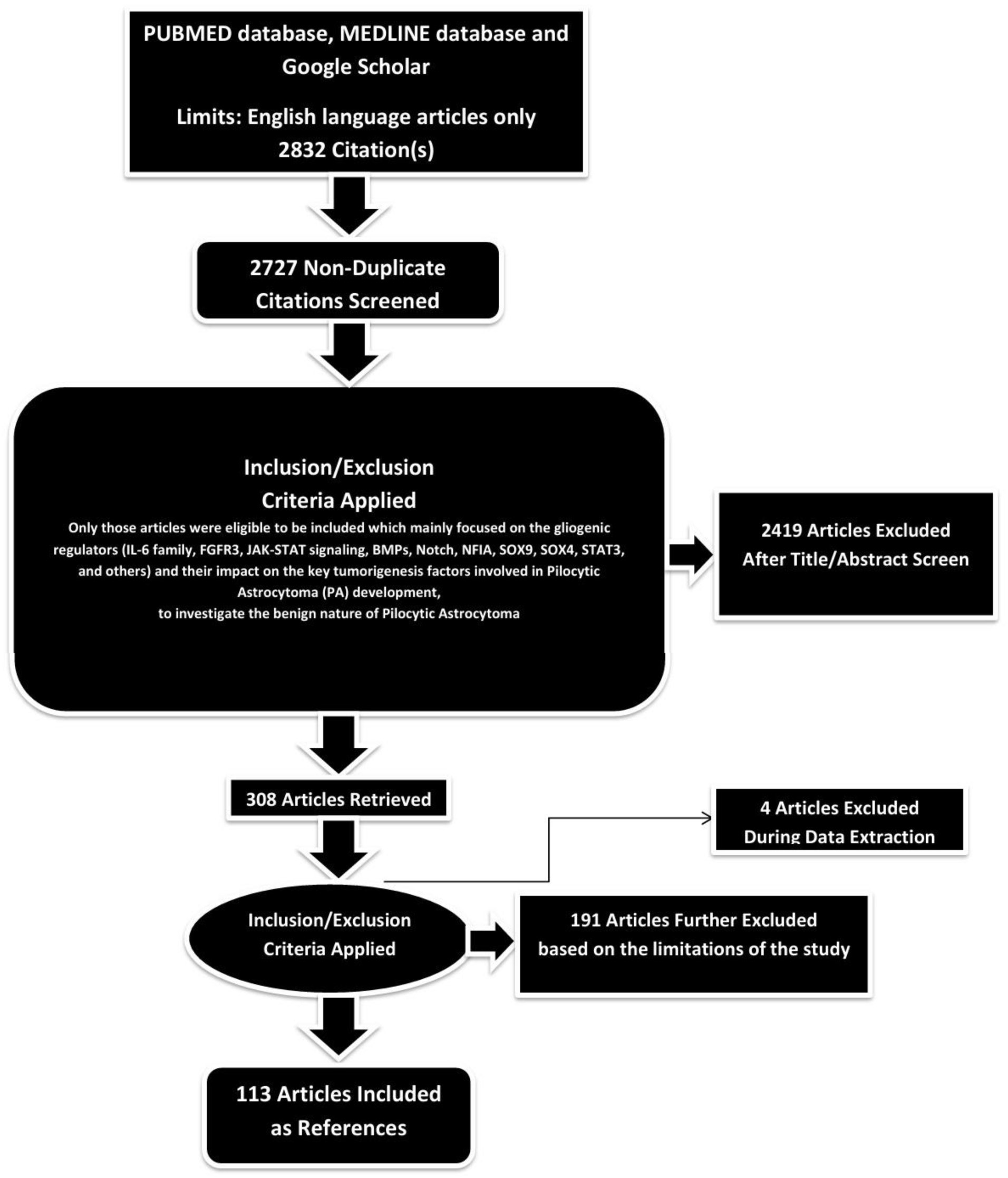
PRISMA FLOW DIAGRAM: This figure represents graphically the flow of citations in the study.

### Assessment of Article Quality and Potential Biases

Ensuring the quality and minimizing potential biases of the selected articles were crucial aspects to guarantee the rigor and reliability of the research findings.

### Quality Assessment

The initial step in quality assessment involved evaluating the methodological rigor of the selected articles. This included a thorough examination of the study design, data collection methods, and analyses conducted. The significance of the study’s findings was weighed based on the quality of the evidence presented. Articles demonstrating sound methodology— such as well-designed studies, controlled variables, and scientifically robust data—were considered of higher quality. Methodological rigor served as a significant indicator of quality.

### Potential Biases Assessment

- Publication Bias: To address the potential for publication bias, a comprehensive search strategy was adopted to include a balanced representation of both positive and negative results, incorporating a wide range of published articles from databases like Google Scholar.
- Selection Bias: Predefined and transparent inclusion criteria were applied to minimize subjectivity in the selection process.

Articles were chosen based on their relevance to the study’s objectives, adhering strictly to these criteria. This approach reduced the risk of subjectivity and ensured that the selection process was objective and consistent.

- Reporting Bias: To mitigate reporting bias, articles were checked for inconsistencies or missing data. Multiple detailed reviews of the methodologies and results were conducted for all selected articles to identify and address any reporting bias.

By including high-quality studies and thoroughly assessing potential biases, this study aimed to provide a robust foundation for the results and conclusions presented.

### Language and Publication Restrictions

We restricted our selection to publications in the English language. There were no limitations imposed on the date of publication. Unpublished studies were not included in our analysis.

By integrating insights from gliogenic regulators, transcription factors, and signaling pathways, this study aims to advance the understanding of Pilocytic Astrocytoma development and progression.

## Results

A total of 2832 articles were identified using database searching, and 2727 were recorded after duplicates removal. 2419 were excluded after screening of title/abstract, 191 were finally excluded, and 4 articles were excluded during data extraction. These exclusions were primarily due to factors such as non-conformity with the study focus, insufficient methodological rigor, or data that did not align with our research question. Finally, 113 articles were included as references.

Astrocytomas are a type of glioma that arise from astrocytes, a subtype of glial cells in the central nervous system (CNS). Astrocytes play crucial roles in neuronal support, blood-brain barrier maintenance, and response to injury. Because of their functional plasticity and proliferative potential, astrocytes or their progenitors are widely considered the cells of origin in astrocytomas [6]. Several lines of evidence support the role of glial cells as the origin of astrocytomas. Genetic and molecular studies have identified mutations in TP53, IDH1/2, ATRX, and PTEN in astrocytomas, particularly in astrocyte-lineage cells. Glioblastoma stem-like cells (GSCs), which exhibit astrocytic properties, further support the notion that astrocytes or glial progenitors contribute to tumor initiation. Experimental models also reinforce this hypothesis, as induced mutations in astrocytes and glial progenitor cells have been shown to lead to astrocytoma development in mouse models, such as the GFAP-Cre and Nestin-Cre models. Additionally, astrocytomas express GFAP (Glial Fibrillary Acidic Protein), a key astrocytic marker, further linking these tumors to an astrocytic origin. Recent studies suggest that neural stem cells (NSCs) and glial progenitors may also serve as the cells of origin, particularly in high-grade astrocytomas like glioblastomas [7, 8].

Despite strong evidence implicating astrocytes and glial progenitors as the primary cells of origin, there is contrasting data suggesting that astrocytes may not be the sole source. While mature astrocytes can dedifferentiate and contribute to tumor formation, neural stem/progenitor cells (NSPCs) may also play a significant role, particularly in aggressive astrocytomas. Some oligodendrocyte precursor cells (OPCs) have also been shown to exhibit transformation potential, indicating that astrocytomas may have a heterogeneous origin. Overall, the strongest evidence supports glial cells—specifically astrocytes and glial progenitors—as the primary cells of origin for astrocytomas. However, neural stem cells (NSCs) and other progenitor populations may also contribute, particularly in higher-grade tumors [9, 10].

### Pilocytic Astrocytoma

Pilocytic Astrocytoma (PA) is primarily driven by genetic alterations affecting the MAPK (Mitogen-Activated Protein Kinase) pathway, with BRAF mutations and fusions being the most common. The most frequent genetic event in PA is the BRAF-KIAA1549 fusion, which leads to constitutive activation of the MAPK pathway. Some cases also exhibit the BRAF V600E mutation, though this is less common in PA compared to higher-grade gliomas. Other genes implicated in PA include NF1, a tumor suppressor gene that negatively regulates RAS signaling and is frequently mutated in optic pathway PAs, particularly in patients with Neurofibromatosis type 1 (NF1). FGFR1 mutations or duplications can also lead to MAPK activation, while PTEN loss, though rare in PA, can contribute to tumor progression. TP53 mutations are uncommon but may be involved in rare cases with malignant transformation [11].

Several transcription factors play crucial roles in astrocytic differentiation, tumor cell proliferation, and MAPK-mediated oncogenesis in PA. SOX10 regulates astrocyte and oligodendrocyte lineage differentiation and is often upregulated in NF1-associated PAs. The Nuclear Factor I (NFI) family, including NFIA, NFIB, and NFIX, controls glial cell development and is expressed in PA cells. STAT3, activated by growth factors and cytokines, contributes to gliogenesis and astrocytoma proliferation. PAX6 is involved in astrocyte lineage differentiation and tumor cell proliferation, while AP-1 (c-Fos/c-Jun complex) is activated through MAPK signaling, promoting tumor growth. Olig2, typically associated with oligodendrocytes, is also sometimes expressed in Pas [12].

Pilocytic Astrocytoma is primarily a MAPK pathway-driven tumor, but additional pathways may contribute to its development and progression. The MAPK/ERK pathway serves as the central oncogenic driver in PA, with BRAF fusions and mutations leading to constitutive ERK activation, promoting proliferation and survival. The PI3K/AKT/mTOR pathway, though primarily involved in high-grade gliomas, interacts with the MAPK pathway and may influence PA development. The JAK/STAT pathway is upregulated in some PAs, particularly through STAT3 activation, influencing astrocytic differentiation and immune response.

The Hedgehog (SHH) pathway, known for its role in glial differentiation, may contribute to PA growth in specific CNS locations. The Notch pathway, which regulates neural stem cell fate, may also play a role in the maintenance of PA cells [13].

Pilocytic Astrocytoma is fundamentally a MAPK-driven tumor, with BRAF fusions and mutations as the predominant oncogenic events. Transcription factors such as SOX10, PAX6, and STAT3 contribute to astrocytic differentiation and tumor cell survival, while the MAPK/ERK pathway remains the dominant signaling cascade. However, interactions with the PI3K, JAK/STAT, and Hedgehog pathways may also influence tumor progression, highlighting the complexity of PA development [14].

### Gliogenic Framework to investigate PA Tumorigenesis

This gliogenic framework is being used to investigate the benign nature of Pilocytic Astrocytoma (PA) development by analyzing key gliogenic genes and signaling pathways that also play roles in glioblastoma (GBM) oncogenesis. Several gliogenic genes have the ability to control oncogenesis in glioblastoma cells, including p300, BMP, PAX6, HOPX, NRSF/REST, LIF, and TGF-β. Among these, PAX6 is particularly notable for its anti-GBM effects, potentially overriding glioblastoma progression, while HOPX exhibits tumor-suppressive and differentiation-promoting properties. NRSF/REST, although primarily astrocytogenic, can also play an oncogenic role in GBM [15]. Some genes and pathways that contribute to gliogenesis also have oncogenic roles in GBM, such as IL-6, FGFR3, JAK-STAT, STAT3, S100, Hey1, HES1, DTX, NF-κB, Neuregulin-1, MAPK, MEK, E2F, TCFL2, NFIX, Ephrins (EFNB1), and Netrins (NTN). These factors are implicated in glioblastoma development but are being examined in PA to determine whether their activity is similarly oncogenic or instead contributes to a more regulated and benign gliogenic process.

Additionally, key genes that contribute to gliogenesis but also play a role in maintaining stemness in GBM include Notch, Sox9, Sox4, and SHH. While these genes are involved in normal gliogenic processes, their overactivation in GBM contributes to cancer stem cell maintenance and tumor progression. Other stemness-related genes such as Nanog, Oct4, and FGF2 are also upregulated in GBM, further promoting oncogenesis. This study looks into how gliogenic regulators play key roles in the onset and development of Pilocytic Astrocytoma and why it remains benign in nature instead of becoming malignant [16, 17].

### Investigating how Gliogenic Regulators contribute to PA Tumorigenesis

#### Impact of Gliogenic Regulators on Key Genetic Alterations in Pilocytic Astrocytoma: BRAF, NF1, FGFR1, PTEN, TP53

##### 1. BRAF

Pilocytic Astrocytoma (PA) is primarily driven by BRAF mutations, particularly BRAF-KIAA1549 fusion, leading to constitutive activation of the MAPK/ERK signaling pathway. Despite this oncogenic activation, PA remains benign rather than progressing into a malignant glioblastoma (GBM). The Gliogenic Framework helps dissect how gliogenic genes, transcription factors (TFs), and signaling pathways contribute to PA’s initiation while maintaining its non-malignant nature [18].

Several key gliogenic genes and pathways influence BRAF-driven tumorigenesis in PA. Among these, some contribute to gliogenesis while acting as tumor suppressors, whereas others, commonly oncogenic in GBM, remain in a restricted activation state, preventing malignancy. PAX6, a known tumor suppressor, plays a critical role in maintaining PA’s benign nature. In PA, PAX6 promotes astrocytic differentiation and inhibits excessive proliferation, counteracting MAPK-driven tumorigenesis. In contrast, GBM frequently exhibits PAX6 downregulation, contributing to an undifferentiated, invasive phenotype. HOPX is another tumor suppressive gene involved in differentiation. In PA, HOPX helps maintain astrocytic identity, limiting uncontrolled proliferation. In GBM, however, HOPX is often downregulated, leading to dedifferentiation and heightened tumor aggressiveness [19].

NRSF/REST, which is primarily associated with gliogenesis, plays a dual role in PA. While REST supports astrocytic differentiation, preventing dedifferentiation into a more aggressive phenotype, it also exhibits oncogenic potential in GBM when overactivated, contributing to cancer stemness and therapy resistance. TGF-β signaling is active in PA but remains restricted to differentiation-promoting functions, ensuring that glial lineage commitment is maintained.

Unlike in GBM, where TGF-β signaling is hijacked for immune suppression, invasion, and stemness maintenance, PA retains a more controlled TGF-β response that reinforces its benign state [20].

While pathways like STAT3 and JAK-STAT signaling are activated in PA, their roles are significantly restricted compared to GBM. In PA, STAT3 is modestly active, possibly supporting proliferation, but it lacks the hyperactivation necessary for promoting glioblastoma-like stemness and invasion. Similarly, S100, HES1, HEY1, and NF-κB, which contribute to gliogenesis and glial identity, do not reach levels of activation seen in GBM. In PA, their expression supports glial differentiation and localized tumor growth rather than promoting an invasive phenotype. In contrast, in GBM, these pathways drive aggressive tumor expansion and therapy resistance. Neuregulin-1, MAPK, MEK, and E2F are also activated in PA due to BRAF-driven MAPK signaling. However, their activation does not trigger the secondary oncogenic programs necessary for malignancy, as seen in GBM. This is likely due to the absence of additional genetic hits, such as TP53 loss or PTEN mutations, which are required for full transformation into high-grade gliomas.

A major reason why PA remains benign is the lack of full activation of gliogenic pathways associated with cancer stemness. Notch signaling, which plays a key role in gliogenesis, is moderately active in PA, ensuring differentiation toward a glial phenotype. However, it does not reach the levels observed in GBM, where hyperactive Notch drives cancer stem cell (CSC) self-renewal and therapy resistance. SOX9 and SOX4, which regulate gliogenic differentiation and neural progenitor fate, are expressed in PA. However, their expression remains below the threshold required for driving dedifferentiation into a GBM-like state. In contrast, GBM hijacks SOX9 to maintain stemness and tumor plasticity. SHH (Sonic Hedgehog signaling) is involved in gliogenesis and is partially active in PA, supporting cellular differentiation. However, its signaling does not reach oncogenic levels, preventing the acquisition of cancer stem-like properties. In GBM, hyperactive SHH contributes to tumor stemness and therapy resistance [21]. Despite oncogenic BRAF activation, PA does not acquire the aggressive features of GBM due to several constraints imposed by gliogenic pathways. The presence of PAX6, HOPX, REST, and controlled TGF-β signaling ensures differentiation and prevents dedifferentiation into an invasive phenotype. These factors, when lost or dysregulated in GBM, contribute to its aggressive behavior. Unlike GBM, PA lacks key mutations such as TP53 loss, PTEN inactivation, or EGFR amplification, which are necessary for high-grade transformation. While MAPK activation is a hallmark of PA, it is insufficient alone to drive full malignancy. Notch, SOX9, and SHH remain below the threshold required for the acquisition of cancer stem-like properties. In GBM, these pathways are hijacked to sustain tumor plasticity and therapeutic resistance, leading to aggressive malignancy [22]. PA does not exhibit high levels of invasion-associated factors such as MMPs,

VEGF, or epithelial-mesenchymal transition (EMT) markers, which are upregulated in GBM to facilitate tumor spread. Applying the Gliogenic Framework to BRAF-mutated PA reveals a delicate balance between gliogenesis and oncogenesis that prevents malignant transformation. Gliogenic tumor suppressors like PAX6, HOPX, and REST maintain differentiation, while oncogenic pathways like STAT3, Notch, and SHH remain partially active, preventing PA from acquiring stem-like properties. Additionally, PA lacks the secondary genetic alterations necessary for transformation into a more aggressive phenotype, unlike GBM, where these alterations drive stemness, invasion, and therapy resistance. Thus, while BRAF activation initiates tumor formation in PA, the interplay of gliogenic factors ensures that PA remains a benign, localized tumor rather than progressing to glioblastoma [23].

##### 2. NF1

Pilocytic Astrocytoma (PA) is commonly associated with alterations in the MAPK/ERK signaling pathway, and one such alteration involves Neurofibromin 1 (NF1), a tumor suppressor gene. NF1 mutations result in constitutive activation of RAS-MAPK signaling, driving tumor formation. However, unlike Glioblastoma (GBM), PA remains benign and localized, suggesting the presence of regulatory mechanisms that prevent full malignant transformation. By applying the Gliogenic Framework, we can examine how gliogenic genes, transcription factors (TFs), and signaling pathways interact with NF1-driven tumorigenesis and contribute to PA’s benign nature [24].

NF1 encodes Neurofibromin, a negative regulator of the RAS-MAPK pathway. Its loss leads to persistent RAS activation, promoting proliferation and glial differentiation defects. However, despite this oncogenic drive, PA does not progress into a high-grade, invasive glioma [25].

Several gliogenic genes play a crucial role in regulating oncogenesis in GBM while restricting malignancy in PA. PAX6 is highly expressed in PA, functioning as a tumor suppressor by promoting astrocytic differentiation and restraining excessive proliferation. In contrast, PAX6 loss in GBM contributes to tumor dedifferentiation and invasiveness. In PA, PAX6 counterbalances NF1 loss, ensuring that astrocytes maintain their differentiated state rather than acquiring stem-like characteristics. HOPX, a tumor-suppressive differentiation factor, is upregulated in PA, facilitating astrocyte maturation and limiting dedifferentiation into a more proliferative state. In GBM, however, HOPX downregulation is associated with poor differentiation and higher tumor plasticity. The sustained activity of HOPX in PA helps prevent malignant transformation. REST regulates astrocytic differentiation by suppressing neuronal gene expression in glial precursors. In PA, moderate REST activity helps maintain a glial phenotype, reinforcing the tumor’s benign nature, whereas REST overactivation in GBM contributes to tumor dedifferentiation and cancer stemness. TGF-β signaling, while supporting gliogenesis, does not reach oncogenic levels in PA. In GBM, TGF-β is frequently hijacked for immune suppression, epithelial-mesenchymal transition (EMT), and invasion, but in PA, it remains restricted to glial differentiation, preventing malignancy [26].

Some gliogenic genes exhibit oncogenic roles in GBM but have limited contributions to PA malignancy. NF1 loss leads to mild activation of STAT3, which can support gliogenesis and localized proliferation. However, STAT3 hyperactivation in GBM promotes stemness, invasion, and therapy resistance. The controlled activation of STAT3 in PA prevents aggressive growth while supporting gliogenesis. Gliogenic regulators such as S100, HES1, HEY1, and NF-κB support astrocytic differentiation in PA but remain below oncogenic levels. In GBM, hyperactive NF-κB and HES1 drive tumor invasion and resistance to apoptosis, whereas in PA, their restricted expression prevents excessive proliferation and invasion. Due to NF1 inactivation, RAS-MAPK signaling remains persistently active in PA, driving low-grade tumor formation. However, the absence of secondary mutations, such as TP53 or PTEN loss, prevents full malignant transformation, differentiating PA from GBM [27]. In PA, Notch activation supports gliogenesis, but it does not reach the high levels required for maintaining glioblastoma-like cancer stem cells (CSCs). In GBM, Notch signaling is hijacked to sustain tumor plasticity, therapy resistance, and invasion, whereas in PA, it remains restricted to normal gliogenic functions. SOX9 and SOX4 regulate gliogenesis, but their expression in PA remains below oncogenic levels, ensuring differentiation rather than dedifferentiation. In GBM, SOX9 is a key player in maintaining CSCs, but in PA, it primarily contributes to glial lineage commitment. SHH (Sonic Hedgehog) signaling is moderately active in PA, supporting gliogenesis but not cancer stemness. In GBM, SHH hyperactivation contributes to stemness, invasion, and therapy resistance, whereas PA’s SHH activity remains below the threshold required for malignancy [28].

Despite NF1 loss and constitutive MAPK activation, PA does not transition into GBM due to multiple regulatory constraints imposed by gliogenic pathways. Tumor suppressor gliogenic genes such as PAX6, HOPX, and REST remain active, ensuring differentiation and preventing tumor progression. In contrast, GBM exhibits PAX6 loss and REST hyperactivation, which drive dedifferentiation and plasticity. Additionally, PA does not acquire secondary mutations in TP53, PTEN, or EGFR, which are critical for malignant transformation. MAPK activation alone is insufficient for GBM-like progression without these secondary genetic alterations. Furthermore, while Notch, SOX9, and SHH contribute to gliogenesis, their activation remains below the threshold for promoting CSC maintenance. In GBM, uncontrolled Notch, SHH, and SOX9 activation sustains cancer stem cells and leads to aggressive malignancy. Another key difference is localized growth without invasion—PA does not exhibit high levels of invasion-associated genes such as MMPs, VEGF, or EMT markers. In contrast, GBM hijacks angiogenic and invasive programs, allowing widespread tumor infiltration.

The Gliogenic Framework provides a mechanistic explanation for why NF1-driven PA remains benign despite MAPK pathway activation. The presence of differentiation-promoting factors such as PAX6, HOPX, and REST, restricted Notch/SHH activity, and the absence of secondary oncogenic hits prevent malignant transformation. Thus, while NF1 loss initiates glioma formation in PA, the interplay of gliogenic regulators ensures it remains a well-differentiated, benign tumor rather than progressing to GBM [29].

##### 3. FGFR1

Pilocytic Astrocytoma (PA) is a low-grade glioma primarily driven by alterations in the MAPK/ERK pathway, with FGFR1 (Fibroblast Growth Factor Receptor 1) mutations playing a significant role. FGFR1 alterations, including activating mutations and duplications, contribute to PA initiation and progression. However, unlike glioblastoma (GBM), PA remains benign and localized despite FGFR1-driven oncogenic signaling. By applying the Gliogenic Framework, we can investigate how gliogenic genes, transcription factors (TFs), and signaling pathways interact with FGFR1-driven tumorigenesis, regulating tumor behavior and preventing malignancy [30].

FGFR1 is a key regulator of gliogenesis and neurodevelopment, playing a central role in astrocyte differentiation and proliferation. In PA, activating FGFR1 mutations lead to sustained MAPK signaling, which promotes low-grade tumor growth but lacks the additional drivers required for malignant transformation. FGFR1 mutations in PA are characterized by constitutive activation of the MAPK/ERK pathway, driving astrocytic proliferation, increased FGFR1-FRS2 signaling, leading to enhanced glial differentiation, and the absence of secondary mutations (e.g., PTEN loss, TP53 mutations), preventing transformation into GBM [31].

Gliogenic genes play a crucial role in restricting malignancy in PA while regulating oncogenesis in GBM. PAX6 remains active in PA, ensuring astrocytic differentiation and growth control, whereas GBM exhibits PAX6 loss, leading to dedifferentiation and increased tumor plasticity. PAX6 interacts with FGFR1, limiting its oncogenic potential by promoting differentiation rather than proliferation. HOPX is upregulated in PA, preventing excessive proliferation despite FGFR1 activation. In contrast, GBM shows HOPX downregulation, which is linked to enhanced tumor plasticity, while its presence in PA reinforces a differentiated astrocytic phenotype. NRSF/REST activity in PA restricts neuronal gene expression and maintains astrocytic identity, whereas in GBM, REST overactivation drives dedifferentiation, stemness, and therapy resistance. TGF-β contributes to gliogenesis and cell differentiation in PA but does not reach the pro-oncogenic levels observed in GBM, where it drives immune suppression, EMT, and invasion. Certain gliogenic genes have oncogenic roles in GBM but limited contribution to PA malignancy. STAT3 is moderately active in PA, contributing to gliogenesis but remaining below oncogenic levels, whereas in GBM, STAT3 hyperactivation supports glioblastoma stem cells (GSCs), invasion, and resistance to apoptosis. S100, HES1, HEY1, and NF-κB support astrocyte differentiation in PA but remain below oncogenic thresholds. In GBM, hyperactivation of these factors drives therapy resistance and invasion, but in PA, their expression remains controlled. FGFR1-driven PA exhibits sustained MAPK signaling, supporting proliferation but not triggering malignant transformation, as the absence of TP53, PTEN mutations, and additional oncogenic hits differentiates PA from GBM [32].

Some gliogenic genes that maintain stemness in GBM are not fully activated in PA. Notch signaling supports gliogenesis in PA but does not reach levels required for sustaining GBM-like cancer stem cells (CSCs). In GBM, Notch hyperactivation contributes to therapy resistance, tumor plasticity, and invasion, whereas in PA, it remains controlled. SOX9 and SOX4 regulate gliogenesis in PA but do not drive dedifferentiation. In GBM, SOX9 overexpression maintains GSCs and enhances tumor aggressiveness, but in PA, it primarily functions in glial lineage specification. SHH (Sonic Hedgehog Signaling) is active in PA but at a restricted level, promoting differentiation rather than cancer stemness. In GBM, uncontrolled SHH signaling supports tumor plasticity and therapy resistance, which is absent in PA.

Despite FGFR1 activation and sustained MAPK signaling, PA remains benign and localized due to multiple regulatory constraints imposed by gliogenic pathways. The presence of tumor suppressor gliogenic genes such as PAX6, HOPX, and REST ensures differentiation and growth control. In GBM, PAX6 loss and REST hyperactivation drive dedifferentiation and plasticity, leading to malignancy. Additionally, PA lacks secondary mutations such as TP53, PTEN loss, and EGFR amplification, which are essential for full malignant transformation. MAPK activation alone does not induce GBM-like behavior without these co-occurring alterations. Restricted activation of cancer stemness pathways ensures that Notch, SOX9, and SHH signaling remain within gliogenic levels in PA rather than promoting CSC maintenance. In GBM, Notch, SOX9, and SHH hyperactivation contribute to CSC formation, therapy resistance, and tumor progression. Furthermore, PA lacks the pro-invasive and angiogenic programs that define GBM, as MMPs, VEGF, and EMT markers remain inactive, preventing infiltration into the surrounding brain tissue [33].

The Gliogenic Framework provides a mechanistic understanding of why FGFR1-driven PA remains benign despite MAPK pathway activation. The presence of differentiation-promoting gliogenic regulators such as PAX6, HOPX, and REST, restricted Notch/SHH activity, and the absence of secondary oncogenic hits prevent full malignant transformation. Thus, while FGFR1 mutations drive glioma formation in PA, gliogenic pathways ensure it remains a well-differentiated, benign tumor rather than progressing to GBM [34, 35].

##### 4. PTEN

PTEN (Phosphatase and Tensin Homolog), a critical tumor suppressor, plays a major role in regulating gliogenic pathways, cell cycle progression, and PI3K/AKT/mTOR signaling. In PA, PTEN expression is usually retained, contrasting with GBM, where its loss is a key driver of malignancy. By applying the Gliogenic Framework, we can investigate how gliogenic transcription factors (TFs), signaling pathways, and molecular regulators interact with PTEN in PA, restricting malignant transformation and maintaining the tumor’s benign nature [36].

PTEN negatively regulates the PI3K/AKT/mTOR pathway, preventing uncontrolled proliferation and promoting differentiation in astrocytes. In PA, PTEN function remains intact, preventing excessive tumor growth and invasion despite the presence of MAPK pathway activation. Preserved PTEN activity keeps PI3K/AKT signaling in check, preventing uncontrolled proliferation. The absence of PTEN loss restricts tumor plasticity and invasion, while PTEN ensures a differentiated astrocytic phenotype, contributing to PA’s benign behavior. In contrast, GBM exhibits frequent PTEN loss, leading to AKT hyperactivation, increased proliferation, and therapy resistance. Dysregulated cell cycle checkpoints allow aggressive growth, and enhanced glioblastoma stem cell (GSC) maintenance promotes tumor heterogeneity [37].

Gliogenic genes play a crucial role in PTEN-driven PA by restricting malignancy. PAX6 remains active in PA, maintaining differentiation and limiting proliferation, whereas in GBM, PTEN loss leads to PAX6 suppression, enabling tumor dedifferentiation and plasticity. HOPX, a differentiation-promoting factor, is retained in PA, reinforcing astrocyte differentiation and preventing malignant transformation, but it is lost in GBM, contributing to tumor stemness and resistance to differentiation signals. NRSF/REST maintains moderate activity in PA, ensuring neural differentiation and preventing stem-like properties, while in GBM, REST is hyperactivated following PTEN loss, promoting stemness and therapy resistance. Controlled TGF-β activity in PA promotes gliogenesis without triggering mesenchymal transformation, whereas in GBM, PTEN loss enhances TGF-β signaling, leading to EMT, invasion, and therapy resistance [38]. Some gliogenic genes have oncogenic roles in GBM but limited contributions to PA malignancy. STAT3 is moderately active in PA, supporting astrocyte differentiation, while in GBM, PTEN loss hyperactivates STAT3, promoting glioblastoma stem cell survival. S100 and NF-κB are active in PA but do not reach levels required for oncogenesis, whereas in GBM, PTEN loss fuels NF-κB and HES1 hyperactivation, driving invasion and therapy resistance. MAPK activation alone is not sufficient to drive PA toward malignancy, while GBM requires PTEN loss for full malignant transformation via hyperactive E2F-driven proliferation. Other gliogenic genes that maintain stemness in GBM are not fully activated in PA. Notch signaling remains within gliogenic levels in PA, preventing dedifferentiation, while in GBM, PTEN loss hyperactivates Notch, enabling glioblastoma stem cell formation. SOX9 remains balanced in PA, ensuring a differentiated state, but in GBM, PTEN loss leads to SOX9 upregulation, maintaining cancer stem-like properties. SHH (Sonic Hedgehog signaling) is active in PA but does not drive uncontrolled proliferation, whereas in GBM, PTEN loss promotes SHH hyperactivation, supporting stem-like trait [39].

Several factors explain why PTEN-expressing Pilocytic Astrocytoma remains benign instead of progressing to GBM. Retention of PTEN suppresses malignancy by maintaining PI3K/AKT control, preventing GBM-like aggressive proliferation, and PA lacks the dedifferentiation signals seen in GBM, maintaining a stable, localized tumor. The presence of tumor suppressor gliogenic genes such as PAX6, HOPX, and REST ensures astrocyte differentiation and growth control, while in GBM, PTEN loss suppresses PAX6 and HOPX, promoting tumor dedifferentiation. Restricted activation of cancer stemness pathways keeps Notch, SOX9, and SHH signaling within gliogenic levels in PA, whereas in GBM, PTEN loss hyperactivates these pathways, driving glioblastoma stem cell maintenance. Furthermore, PA lacks the invasive potential seen in GBM due to PTEN retention, and pro-invasive markers such as VEGF, MMPs, and EMT remain inactive in PA. The Gliogenic Framework provides a mechanistic explanation for why PTEN-retaining PA remains a benign, well-differentiated glioma. Unlike GBM, which loses PTEN and undergoes malignant transformation, PA retains its tumor suppressive mechanisms, restricting PI3K/AKT-driven oncogenesis, maintaining differentiation, and preventing invasion. Thus, while PA exhibits oncogenic MAPK pathway activation, gliogenic regulators ensure it remains a well-differentiated, non-invasive tumor rather than progressing to GBM [40, 41].

##### 5. TP53

While TP53 mutations are rare in PA, its functional role in gliogenic regulation is crucial for understanding why PA remains benign rather than progressing to malignancy. By applying the Gliogenic Framework, we can analyze how gliogenic transcription factors (TFs), signaling pathways, and tumor suppressors interact with TP53 in PA, regulating its growth, differentiation, and non-invasive nature [42].

TP53 is a critical tumor suppressor, regulating cell cycle arrest, apoptosis, and differentiation in glial progenitors. In PA, TP53 is generally wild-type and functional, preventing excessive proliferation and maintaining cellular differentiation. Wild-type TP53 prevents uncontrolled proliferation and genomic instability, supports astrocytic differentiation by regulating gliogenic genes, and ensures cellular senescence mechanisms are intact, preventing malignant transformation. In contrast, GBM exhibits frequent TP53 mutations, leading to uncontrolled proliferation due to the loss of cell cycle checkpoints, enhanced resistance to apoptosis, promoting therapy resistance, and genomic instability, allowing accumulation of additional oncogenic mutations [43].

Gliogenic genes play a crucial role in restricting malignancy in PA while regulating oncogenesis in GBM. PAX6 remains active in PA, promoting differentiation and suppressing proliferation, whereas in GBM, TP53 loss leads to PAX6 downregulation, enabling tumor dedifferentiation and plasticity. HOPX is retained in PA, reinforcing astrocytic differentiation and preventing malignant transformation, but it is lost in GBM, contributing to stem-like properties and uncontrolled growth. NRSF/REST exhibits moderate activity in PA, maintaining differentiation and preventing stem-like traits, whereas in GBM, TP53 loss leads to REST overactivation, increasing tumor plasticity and therapy resistance. TGF-β signaling remains controlled in PA, supporting differentiation without promoting invasion, whereas in GBM, TP53 loss allows hyperactivation of TGF-β, driving epithelial-to-mesenchymal transition (EMT) and invasion. Certain gliogenic genes exhibit oncogenic roles in GBM but have a limited contribution to PA malignancy. STAT3 is moderately active in PA, supporting astrocyte differentiation, while in GBM, TP53 loss leads to STAT3 hyperactivation, supporting glioblastoma stem cell (GSC) maintenance. Factors such as S100, HES1, HEY1, and NF-κB remain within gliogenic ranges in PA, preventing excessive tumor growth, whereas in GBM, TP53 loss deregulates these pathways, driving inflammation and invasion. MAPK activation alone does not induce full malignancy in PA, whereas GBM requires TP53 loss to enable E2F-driven proliferation and tumor progression [44].

Additionally, some gliogenic genes maintain stemness in GBM but are not fully activated in PA. Notch signaling remains within gliogenic levels in PA, preventing dedifferentiation, whereas in GBM, TP53 loss hyperactivates Notch, maintaining glioblastoma stem cells (GSCs). SOX9 remains controlled in PA, maintaining differentiation, while in GBM, TP53 loss leads to SOX9 upregulation, supporting cancer stem-like properties. SHH signaling is active in PA but does not drive uncontrolled proliferation, whereas in GBM, TP53 loss promotes SHH hyperactivation, supporting tumor progression [45].

TP53-wildtype PA remains benign instead of progressing to GBM due to several key factors. Retention of TP53 suppresses malignancy by maintaining cell cycle checkpoints and preventing uncontrolled proliferation. PA lacks the dedifferentiation signals seen in GBM, ensuring a stable, localized tumor. The presence of tumor suppressor gliogenic genes such as PAX6, HOPX, and REST ensures astrocyte differentiation and restricts growth, while in GBM, TP53 loss suppresses these genes, enabling tumor dedifferentiation. Cancer stemness pathways, including Notch, SOX9, and SHH signaling, remain within gliogenic levels in PA, whereas in GBM, TP53 loss hyperactivates these pathways, driving glioblastoma stem cell formation. PA lacks the invasive potential seen in GBM due to TP53 retention, and pro-invasive markers such as VEGF, MMPs, and EMT remain inactive in PA. The Gliogenic Framework provides a mechanistic explanation for why TP53-wildtype PA remains a benign, well-differentiated glioma. Unlike GBM, which frequently loses TP53 and undergoes malignant transformation, PA retains its tumor-suppressive mechanisms, preventing genomic instability, hyperproliferation, and invasion [46, 47].

#### Impact of Gliogenic Regulators on Transcription Factors (TFs) Implicated in PA Development: SOX10, NFI (Nuclear Factor I Family: NFIA, NFIB, NFIX), STAT3, PAX6, AP-1, Olig2

##### 1. SOX10

The SOX10 transcription factor is primarily known for its role in neural crest-derived glial lineage specification, but its involvement in gliogenesis and tumorigenesis makes it an important player in understanding PA biology. By applying the Gliogenic Framework, we can analyze how gliogenic genes, signaling pathways, and differentiation regulators interact with SOX10 in PA, ultimately influencing why PA remains benign rather than becoming malignant like GBM [48]. SOX10 is a key transcription factor regulating gliogenesis, particularly in oligodendrocyte precursor cells (OPCs) and Schwann cells. While SOX10 is primarily known for driving oligodendrocyte differentiation, it also influences astrocyte differentiation and neural tumor progression, making it a relevant factor in PA tumor biology. SOX10 remains expressed at gliogenic levels in PA but does not drive malignant transformation, regulates differentiation pathways in astrocytes and oligodendrocytes to prevent dedifferentiation, and maintains a balance between gliogenic fate specification and tumor suppressive differentiation programs. In contrast, in GBM, SOX10 function is often altered, promoting glioblastoma stem-like properties, interacting with pro-oncogenic pathways such as Notch and Wnt to enhance tumor plasticity, and contributing to aggressive tumor progression and therapy resistance [49].

Gliogenic genes that regulate SOX10 play a crucial role in restricting malignancy in PA. PAX6 interacts with SOX10 to promote astrocyte differentiation in PA, whereas in GBM, PAX6 loss leads to loss of differentiation control, enabling malignant transformation. HOPX restricts SOX10 activity to a pro-differentiation state, preventing stem-like features, while in GBM, HOPX is downregulated, allowing SOX10 to contribute to tumor plasticity. REST prevents SOX10-driven dedifferentiation in PA, maintaining a benign phenotype, whereas in GBM, REST is often deregulated, allowing SOX10 to promote tumor stemness. TGF-β signaling remains controlled in PA, ensuring that SOX10 supports differentiation rather than proliferation, while in GBM, TGF-β hyperactivation allows SOX10 to contribute to glioblastoma plasticity and invasion [50].

Certain gliogenic genes with oncogenic roles in GBM have a limited contribution to PA malignancy. STAT3 remains at gliogenic levels in PA, supporting differentiation and preventing malignancy, while in GBM, STAT3 hyperactivation allows SOX10 to enhance glioblastoma stem cell (GSC) maintenance. Factors such as S100, HES1, HEY1, and NF-κB are not overactivated in PA, keeping SOX10 within differentiation-associated functions, while in GBM, their overactivation allows SOX10 to contribute to tumor progression. MAPK activation alone in PA does not lead to malignancy because SOX10 remains regulated, whereas in GBM, SOX10 interacts with E2F-driven pathways to promote tumor cell proliferation. Gliogenic genes that maintain stemness in GBM are not fully activated in PA. Notch remains in a controlled state in PA, preventing SOX10-driven dedifferentiation, while in GBM, Notch hyperactivation allows SOX10 to support glioblastoma stem-like states. SOX9 interacts with SOX10 to maintain differentiation in PA, whereas in GBM, SOX9-SOX10 interactions support cancer stem cell properties. SHH remains at normal gliogenic levels in PA, preventing SOX10 from driving tumor progression, while in GBM, SHH hyperactivation supports SOX10-induced glioblastoma plasticity [51].

SOX10-wildtype pilocytic astrocytoma remains benign instead of progressing to GBM due to several key factors. SOX10 maintains a pro-differentiation role in PA, limiting tumorigenesis, whereas in GBM, SOX10 interacts with stemness pathways such as Notch and SHH to drive plasticity. The presence of tumor suppressor gliogenic genes such as PAX6, HOPX, and REST ensures differentiation and restricts proliferation, whereas in GBM, their loss allows SOX10 to contribute to tumor progression. SOX10 does not interact with cancer stemness pathways in PA, maintaining a stable tumor phenotype, while in GBM, SOX10 interacts with SOX9, Notch, and SHH to drive malignancy. PA lacks the invasive characteristics seen in GBM due to controlled SOX10 activity, with pro-invasive markers such as VEGF, MMPs, and EMT remaining inactive in PA. The Gliogenic Framework provides a mechanistic explanation for why SOX10-wildtype PA remains a benign, well-differentiated glioma. Unlike GBM, which frequently deregulates SOX10 to support glioblastoma stem-like features, PA retains its tumor suppressive differentiation mechanisms, preventing genomic instability, dedifferentiation, and invasion. Thus, while PA exhibits MAPK pathway activation, gliogenic regulators ensure SOX10 remains a differentiation-supporting factor rather than a driver of malignancy [52, 53].

##### 2. NFI (Nuclear Factor I Family: NFIA, NFIB, NFIX)

The Nuclear Factor I (NFI) transcription factor family—comprising NFIA, NFIB, and NFIX—plays a crucial role in gliogenesis, astrocytic differentiation, and brain tumor biology. By applying the Gliogenic Framework, we can investigate how NFI factors regulate PA initiation and maintenance, preventing its progression into a high-grade, invasive glioma [54].

The NFI family (NFIA, NFIB, and NFIX) regulates gliogenesis by controlling glial progenitor expansion, astrocyte maturation, and neural stem cell (NSC) fate decisions. In PA, these transcription factors play a critical role in maintaining astrocytic differentiation while limiting dedifferentiation into stem-like glioma cells, restricting tumor growth by balancing proliferation and differentiation signals, and interacting with MAPK signaling to support a benign, well-differentiated state. In contrast, glioblastoma (GBM) frequently deregulates NFI factors, promoting tumor plasticity and invasion. In PA, NFIA/NFIB/NFIX expression is controlled, supporting astrocyte differentiation, while in GBM, their expression is altered, contributing to glioblastoma plasticity. The interaction between NFI factors and MAPK signaling supports differentiation without uncontrolled proliferation in PA, whereas in GBM, these interactions facilitate oncogenic pathway activation, leading to tumor growth. Furthermore, while PA restricts dedifferentiation, GBM fosters glioblastoma stem cell properties, and PA suppresses pro-invasive markers, while GBM activates epithelial-to-mesenchymal transition (EMT) and pro-metastatic factors [55].

Several gliogenic genes support NFIA/NFIB/NFIX in restricting malignancy in PA. PAX6 cooperates with NFIA/NFIB/NFIX to promote astrocyte identity, while its loss in GBM removes this restriction, allowing NFIA/NFIB to promote tumor progression. HOPX maintains NFIA-driven astrocyte differentiation, preventing PA from acquiring stem-like features, but in GBM, HOPX loss enables NFIA/NFIX to support glioblastoma plasticity. REST suppresses NFIA-driven glioblastoma stem cell formation in PA, whereas in GBM, REST is downregulated, allowing NFI factors to contribute to tumor progression. TGF-β remains controlled in PA, preventing NFIA/NFIX from driving tumor plasticity, but in GBM, TGF-β hyperactivation allows NFIA/NFIX to enhance invasion and dedifferentiation [56].

Certain gliogenic genes become oncogenic in GBM but remain restricted in PA. STAT3 interacts with NFIA in gliogenesis but does not promote tumorigenesis in PA, while in GBM, the STAT3-NFIA interaction drives glioblastoma stem-like features. S100B, HES1, HEY1, and NF-κB, which regulate proliferation and plasticity, are limited by NFIA/NFIB in PA, whereas in GBM, their activation allows NFI factors to enhance tumor progression. NFIA/NFIB interact with MAPK in PA to support differentiation without uncontrolled proliferation, but in GBM, MAPK-NFI interactions promote aggressive tumor growth. Gliogenic genes that maintain stemness in GBM are not fully activated in PA. Notch signaling, which is crucial in NSC fate, is regulated in PA, preventing NFIA/NFIX from inducing stem-like states, while in GBM, Notch overactivation enables NFI factors to drive glioblastoma plasticity. SOX9 and SOX10 work with NFIA to maintain astrocyte identity in PA, whereas in GBM, SOX9/NFI interactions promote tumor initiation and progression. SHH signaling is limited in PA, ensuring that NFI factors promote differentiation rather than tumor progression, but in GBM, SHH activation allows NFIA/NFIX to contribute to glioblastoma formation.

NFI-wildtype Pilocytic Astrocytoma remains benign instead of progressing to GBM due to multiple factors. NFIA-driven differentiation prevents PA from acquiring invasive, stem-like properties, whereas in GBM, NFI factors interact with oncogenic pathways, driving tumor plasticity. Tumor suppressor gliogenic genes such as PAX6, HOPX, and REST regulate NFIA/NFIB/NFIX, ensuring differentiation and restricting proliferation, but in GBM, their loss allows NFI factors to enhance malignancy. Cancer stemness pathways are restricted in PA, preventing NFI factors from interacting with oncogenic drivers, whereas in GBM, NFIA/NFIX interact with SOX9, Notch, and SHH to drive malignancy. Additionally, PA exhibits localized growth without invasion due to controlled NFIA/NFIX activity, while pro-invasive markers such as VEGF, MMPs, and EMT remain inactive in PA [57].

The Gliogenic Framework provides a mechanistic explanation for why NFI-wildtype PA remains a benign, well-differentiated glioma. Unlike GBM, where NFIA/NFIB/NFIX contribute to glioblastoma plasticity and invasion, PA retains its tumor-suppressive differentiation mechanisms, preventing dedifferentiation, invasion, and high proliferative capacity. While PA exhibits MAPK pathway activation, gliogenic regulators ensure that NFIA/NFIB/NFIX remain differentiation-supporting factors rather than drivers of malignancy [58, 59].

##### 3. STAT3

Signal Transducer and Activator of Transcription 3 (STAT3) is a key regulator of gliogenesis, neural stem cell maintenance, and oncogenic transformation in gliomas. Its role in Pilocytic Astrocytoma (PA) is paradoxical: while STAT3 is required for astrocyte differentiation and survival, its deregulation in high-grade gliomas (e.g., glioblastoma, GBM) promotes tumor plasticity, invasion, and therapy resistance. Applying the Gliogenic Framework allows us to investigate how STAT3 functions in PA development and why PA remains benign instead of progressing into malignant glioma [60].

STAT3 functions as both a pro-differentiation factor in normal gliogenesis, ensuring astrocytes mature correctly, and as a tumor-promoting factor when hyperactivated, supporting glioblastoma stemness, immune evasion, and invasion. In Pilocytic Astrocytoma, STAT3 is involved in differentiation without oncogenic transformation, ensuring that PA remains a slow-growing, localized tumor rather than an invasive malignancy. Compared to GBM, PA exhibits controlled STAT3 activation that supports differentiation, limited interaction with JAK/IL-6 to maintain normal astrocyte fate, restricted stemness promotion to prevent self-renewal, suppression of invasion and epithelial-mesenchymal transition (EMT) induction, and moderate inflammatory signaling that preserves immune homeostasis. In contrast, GBM exhibits hyperactive STAT3 that drives tumor progression, persistent JAK/IL-6 activation that promotes growth, enhanced glioblastoma stem-like properties, activated invasion mechanisms, and elevated inflammatory signaling that supports tumor immune escape [61].

Several gliogenic genes support STAT3 in preventing malignancy in PA. PAX6 inhibits hyperactivation of STAT3, maintaining a differentiated state, while its loss in GBM enables STAT3 to drive glioblastoma stem cell (GSC) formation. NFIA, NFIB, and NFIX work with STAT3 to ensure astrocyte differentiation in PA, whereas in GBM, these factors cooperate with hyperactivated STAT3 to sustain tumor plasticity. HOPX prevents STAT3 from inducing self-renewal programs in PA, but its loss in GBM enables STAT3-driven oncogenesis. TGF-β signaling remains controlled in PA, preventing STAT3 from promoting malignancy, whereas in GBM, overactivation enhances STAT3-induced mesenchymal transition and invasion [62].

Other gliogenic genes that become oncogenic in GBM but remain restricted in PA include the JAK-STAT3 pathway, which is only moderately and transiently activated in PA but chronically active in GBM, driving tumor progression. Pro-inflammatory cytokines such as IL-6, IL-10, and NF-κB remain low in PA, preventing STAT3-driven glioblastoma plasticity, but are elevated in GBM, amplifying STAT3 activation and malignancy. Stemness regulators like S100B, HES1, and HEY1 are limited in PA, restricting STAT3 function, while their overexpression in GBM promotes tumor dedifferentiation and invasion. STAT3 does not induce VEGF, MMPs, or EMT genes in PA, thereby restricting malignancy, whereas in GBM, STAT3 interacts with hypoxia-induced VEGF and EMT pathways to enhance invasion. Gliogenic genes that maintain stemness in GBM but are not fully activated in PA further reinforce PA’s benign nature. Notch signaling remains balanced in PA, preventing STAT3 from sustaining glioblastoma stemness, whereas its overactivation in GBM enhances STAT3-mediated self-renewal. STAT3 cooperates with SOX9 in PA to ensure proper astrocyte differentiation, while in GBM, SOX9/STAT3 interactions promote glioblastoma initiation. Limited Sonic Hedgehog (SHH) activation in PA prevents STAT3 from promoting dedifferentiation, whereas in GBM, SHH-STAT3 cooperation enhances tumor growth and resistance [63, 64].

STAT3 promotes differentiation rather than oncogenesis in PA by supporting astrocyte maturation rather than tumor plasticity. In contrast, in GBM, STAT3 interacts with oncogenic pathways to sustain glioblastoma growth. The presence of tumor suppressor gliogenic genes such as PAX6, NFIA, NFIB, NFIX, and HOPX restricts STAT3’s oncogenic potential in PA, whereas their loss in GBM allows STAT3 to function as a tumor promoter. Additionally, STAT3 is not fully engaged in self-renewal in PA, while in GBM, it is activated by Notch, SOX9, and SHH, driving malignancy. Finally, PA exhibits localized growth without invasion since STAT3 does not induce VEGF, MMPs, or EMT-related genes, whereas in GBM, STAT3 cooperates with hypoxia pathways to promote angiogenesis and invasion. STAT3 plays a dual role in gliogenesis. In PA, STAT3 maintains astrocyte differentiation, preventing tumor dedifferentiation and invasion, while in GBM, STAT3 hyperactivation interacts with Notch, JAK-STAT, and TGF-β pathways to drive glioblastoma stemness, plasticity, and invasion. The Gliogenic Framework explains why STAT3 remains a differentiation-supporting factor in PA instead of promoting malignancy. Unlike in GBM, where STAT3 is persistently activated, PA retains mechanisms that suppress its tumorigenic potential, ensuring a benign, slow-growing tumor phenotype [65].

##### 4. PAX6

PAX6 (Paired Box 6) is a critical neurogenic transcription factor that plays a key role in glial differentiation, tumor suppression, and lineage specification. While PAX6 loss is associated with high-grade gliomas such as glioblastoma (GBM), it remains highly expressed in Pilocytic Astrocytoma (PA), ensuring the tumor’s benign and well-differentiated nature. Applying the Gliogenic Framework allows us to investigate how PAX6 functions in PA development and why PA remains non-invasive and localized instead of progressing into a malignant glioma [66].

PAX6 functions as both a gliogenic differentiation factor, guiding astrocyte maturation, and a tumor suppressor, inhibiting stemness and tumor plasticity. In Pilocytic Astrocytoma, PAX6 expression remains high, supporting differentiation and preventing oncogenic transformation, whereas in GBM, its loss leads to a loss of lineage identity and tumor plasticity. In PA, PAX6 restricts self-renewal, suppresses invasion and epithelial-to-mesenchymal transition (EMT) induction, and limits angiogenesis by keeping VEGF activation low. Conversely, in GBM, PAX6 loss enables tumor-initiating cells, enhances migration, and promotes aggressive vascular growth. This highlights the role of PAX6 as a gliogenic gatekeeper in PA, maintaining a slow-growing, well-defined tumor [67].

The Gliogenic Framework identifies several gliogenic genes that support PAX6 in preventing malignancy in PA. In PA, PAX6 works with STAT3 to maintain astrocyte differentiation, whereas in GBM, STAT3 activation without PAX6 promotes tumor plasticity. PAX6 enhances the function of NFIA, NFIB, and NFIX to ensure astrocyte differentiation in PA, but in GBM, NFIA can cooperate with tumor-promoting factors to drive plasticity. PAX6 also interacts with HOPX to suppress dedifferentiation in PA, while in GBM, HOPX loss allows cells to re-enter a tumor-initiating state.

Additionally, PAX6 modulates SOX9 and SOX10 to ensure mature glial fate in PA, while in GBM, SOX9/10 overexpression without PAX6 leads to neural crest-like dedifferentiation [68]. In PA, PAX6 suppresses chronic JAK-STAT3 activation, preventing inflammation-induced tumorigenesis, whereas in GBM, loss of PAX6 allows persistent STAT3 signaling, fueling tumor growth. Similarly, the stemness regulators S100B, HES1, and HEY1 have limited activity in PA, ensuring a restricted progenitor-like state, but in GBM, their overexpression supports tumor self-renewal and plasticity. PAX6 also controls TGF-β signaling in PA to prevent tumor cell migration, while in GBM, TGF-β overactivation with low PAX6 induces EMT and invasiveness. Gliogenic genes that maintain stemness in GBM but are not fully activated in PA further support the benign nature of PAX6-wildtype PA. In PA, Notch activity is regulated by PAX6, preventing self-renewal, while in GBM, Notch activation without PAX6 enables glioblastoma stemness. PAX6 also suppresses Sonic Hedgehog (SHH)-driven tumorigenesis in PA, whereas in GBM, SHH signaling cooperates with glioblastoma-initiating factors. Additionally, PAX6 limits VEGF and MMP activity in PA, restricting vascular expansion, while in GBM, its loss allows VEGF-induced tumor invasion [69].

The reason why PAX6-wildtype Pilocytic Astrocytoma remains benign instead of progressing to GBM is that PAX6 restricts tumor plasticity, preventing glioblastoma-like dedifferentiation and maintaining a stable astrocytic phenotype. In GBM, PAX6 loss allows stem-like reprogramming, leading to tumor heterogeneity. The presence of tumor suppressor gliogenic genes such as STAT3, NFIA/NFIB/NFIX, and HOPX, which cooperate with PAX6 to prevent malignant transformation in PA, also plays a crucial role. In GBM, their loss amplifies tumorigenic potential.

Furthermore, PAX6 suppresses oncogenic pathways like JAK-STAT, Notch, and TGF-β in PA, while in GBM, these pathways become hyperactivated due to PAX6 downregulation. The localized growth of PA without invasion is also maintained by PAX6, which prevents VEGF/MMP-driven angiogenesis and invasion, unlike in GBM, where PAX6 loss enables vascular remodeling and EMT-mediated invasion [70].

PAX6 plays a dual role in gliogenesis. In PA, it maintains gliogenic differentiation, preventing tumor dedifferentiation and invasion, while in GBM, its loss enables oncogenic transformation, stem-like plasticity, and tumor invasion. The Gliogenic Framework explains why PAX6 remains a differentiation-supporting factor in PA, ensuring it remains benign and non-invasive. Unlike GBM, where PAX6 is downregulated, PA retains gliogenic mechanisms that suppress its tumorigenic potential, ensuring a well-defined, slow-growing tumor phenotype [71].

##### 5. AP-1

Activator Protein-1 (AP-1) is a transcription factor complex composed of JUN, FOS, ATF, and MAF proteins that plays a dual role in gliogenesis and tumor progression. In Pilocytic Astrocytoma (PA), AP-1 is active in a controlled, differentiation-supporting manner, ensuring the tumor remains benign and non-invasive. However, in glioblastoma (GBM), AP-1 activity is often dysregulated, leading to enhanced proliferation, invasion, and malignant transformation. Using the Gliogenic Framework, we will examine how AP-1 interacts with key gliogenic players in PA and why AP-1-driven signaling remains non-oncogenic in PA but drives malignancy in GBM [72].

In PA, AP-1 (JUN/FOS) activity is controlled by gliogenic regulators such as PAX6, STAT3, and NFIA, ensuring a differentiated glial phenotype. In contrast, in GBM, AP-1 hyperactivation promotes tumor dedifferentiation, invasion, and stemness. The controlled AP-1 activity in PA ensures that glial differentiation is promoted while tumorigenic plasticity is suppressed, explaining its benign nature. AP-1 activity in PA remains moderate and differentiation-supportive, whereas in GBM, hyperactive AP-1 drives proliferation and invasion. AP-1 maintains astrocyte differentiation in PA through its interactions with STAT3, PAX6, and NFIA, but in GBM, loss of these controls leads to tumor plasticity. The oncogenic stress response in PA is transient, while in GBM, AP-1 sustains oncogenic transcription. Furthermore, AP-1 remains non-invasive in PA, whereas in GBM, it induces MMPs and VEGF, promoting tumor progression [73].

Several gliogenic genes play a role in supporting AP-1 in preventing malignancy in PA. PAX6 acts as a tumor suppressor and differentiation regulator, restricting AP-1 activity to differentiation-associated genes in PA. In GBM, downregulation of PAX6 allows AP-1 to activate oncogenic programs. STAT3, which influences glial fate, interacts with AP-1 in PA to promote astrocyte differentiation, but in GBM, persistent STAT3-AP-1 signaling enhances tumor survival and invasion. Astrocytic differentiation regulators such as NFIA, NFIB, and NFIX work alongside AP-1 to maintain a non-proliferative, benign phenotype in PA, but in GBM, their dysregulation allows AP-1 to promote mesenchymal transitions. HOPX, another tumor suppressor, interacts with AP-1 in PA to prevent tumor dedifferentiation, whereas in GBM, the loss of HOPX enables AP-1 to promote glioblastoma plasticity [74].

AP-1’s activation of JAK-STAT3 is transient in PA, preventing tumor progression, while in GBM, sustained JAK-STAT3 activation leads to pro-tumor inflammation. PAX6 limits the interaction between AP-1 and SOX9/SOX10 in PA, restricting progenitor-like states, but in GBM, SOX9/SOX10 are hyperactivated by AP-1, driving dedifferentiation and invasion. AP-1 does not strongly activate MMPs and VEGF in PA, thereby preventing aggressive growth, while in GBM, it enhances VEGF/MMP signaling, promoting vascular expansion and metastasis. Additionally, some gliogenic genes that maintain stemness in GBM are not fully activated in PA. AP-1 does not strongly interact with Notch in PA, limiting tumorigenesis, whereas in GBM, it cooperates with Notch to sustain glioblastoma stemness. AP-1 does not induce SHH-driven tumorigenesis in PA, but in GBM, it amplifies SHH signaling, promoting tumor progression. In PA, TGF-β signaling remains controlled, preventing AP-1-driven invasion, whereas in GBM, AP-1 cooperates with TGF-β to drive epithelial-to-mesenchymal transition (EMT) and glioblastoma metastasis [75].

AP-1 in PA is differentiation-supportive rather than oncogenic due to its regulation by gliogenic factors such as PAX6, STAT3, and NFIA. In GBM, AP-1 hyperactivation bypasses differentiation, leading to increased invasion and malignancy. The suppression of AP-1-driven inflammatory pathways in PA prevents a chronic inflammatory tumor microenvironment, whereas in GBM, persistent AP-1 activation contributes to tumor progression. AP-1 does not strongly activate VEGF, MMPs, or EMT genes in PA, limiting angiogenesis and invasion, while in GBM, AP-1 induces these genes, enhancing tumor progression. In PA, the AP-1-STAT3 interaction maintains glial differentiation, whereas in GBM, its deregulation allows tumor stemness and dedifferentiation.

AP-1 acts as a context-dependent regulator in gliogenesis. In PA, its activity is tightly regulated by PAX6, STAT3, and NFIA, ensuring differentiation and limiting oncogenesis. In GBM, the loss of these regulatory controls allows AP-1 to drive oncogenic transcription, invasion, and tumor dedifferentiation. The Gliogenic Framework explains why AP-1 remains a differentiation-promoting factor in PA, preventing the tumor from becoming invasive or malignant. Unlike GBM, where AP-1 hyperactivation fuels progression, PA retains gliogenic regulators that suppress AP-1-driven tumorigenic plasticity, ensuring it remains benign and slow-growing [76, 77].

##### 6. Olig2

Oligodendrocyte transcription factor 2 (Olig2) is a critical basic helix-loop-helix (bHLH) transcription factor that regulates glial lineage commitment, neural progenitor maintenance, and glioma progression. While Olig2 is a pro-gliogenic factor, its dysregulation in glioblastoma (GBM) leads to increased tumor plasticity, self-renewal, and therapy resistance. In Pilocytic Astrocytoma (PA), Olig2 expression is moderate and controlled, promoting glial differentiation rather than tumorigenesis. Using the Gliogenic Framework, we will analyze why Olig2 does not drive malignancy in PA and how it interacts with other gliogenic regulators to maintain a benign phenotype [78].

In PA, Olig2 promotes astrocyte differentiation while preventing stemness. In contrast, in GBM, Olig2 enhances glioma-initiating cell (GIC) properties, promoting malignancy. Olig2’s controlled activity in PA ensures differentiation over oncogenesis, explaining why PA remains benign. Comparatively, in GBM, hyperactive Olig2 drives tumorigenic plasticity. In PA, Olig2 interacts with gliogenic regulators like PAX6, STAT3, and NFIA to support glial differentiation, while in GBM, these interactions are disrupted, enhancing tumor self-renewal and therapy resistance. Unlike GBM, PA maintains controlled glial commitment, preventing Olig2-mediated progenitor expansion and limiting therapeutic resistance [79].

Several gliogenic genes support Olig2 in preventing malignancy in PA. PAX6 functions as a tumor suppressor and differentiation regulator, restraining Olig2 activity and ensuring astrocyte differentiation. In GBM, PAX6 downregulation allows Olig2 to drive glioma-initiating cell properties. STAT3 directs Olig2 toward differentiation-associated transcription in PA, whereas in GBM, hyperactive STAT3-Olig2 signaling extends tumor plasticity and self-renewal. NFIA, NFIB, and NFIX cooperate with Olig2 to drive astrocyte fate rather than progenitor expansion in PA. In contrast, their loss in GBM allows Olig2 to sustain a progenitor-like state, enhancing malignancy. HOPX, another tumor suppressor, modulates Olig2 activity in PA, preventing dedifferentiation, whereas in GBM, its loss allows Olig2 to drive glioma stem-like features [80].

Some gliogenic genes that become oncogenic in GBM are restricted in PA. Olig2-promoted JAK-STAT3 activation remains transient in PA, preventing prolonged oncogenic signaling, whereas in GBM, persistent JAK-STAT3-Olig2 interaction sustains tumor cell survival and invasion. SOX9 and SOX10, which contribute to stemness and dedifferentiation, are limited by PAX6 in PA, restricting progenitor-like states. In GBM, SOX9 and SOX10 are hyperactivated by Olig2, promoting self-renewal and therapy resistance. Similarly, matrix metalloproteinases (MMPs) and vascular endothelial growth factor (VEGF), which contribute to angiogenesis and tumor invasion, are not strongly activated by Olig2 in PA, limiting tumor vascularization, whereas in GBM, Olig2 enhances their expression, facilitating invasion. Several gliogenic genes that maintain stemness in GBM are not fully activated in PA. Notch signaling, which regulates tumor-initiating cells, does not strongly interact with Olig2 in PA, preventing tumorigenesis, while in GBM, Olig2-Notch interaction promotes glioma-initiating cells. Sonic Hedgehog (SHH) signaling, crucial for neural stem cell maintenance, is not induced by Olig2 in PA, whereas in GBM, Olig2 amplifies SHH signaling, sustaining tumor growth. The TGF-β pathway, which drives epithelial-to-mesenchymal transition (EMT) and invasion, remains controlled in PA, preventing Olig2-driven invasion. However, in GBM, Olig2 cooperates with TGF-β to enhance EMT and glioblastoma metastasis [81].

Olig2 remains within a gliogenic differentiation network controlled by PAX6, STAT3, and NFIA in PA, promoting differentiation rather than stemness. In contrast, in GBM, Olig2 escapes differentiation constraints, driving progenitor-like and tumorigenic states. Gliogenic factors suppress Olig2’s ability to maintain neural progenitor-like states in PA, while in GBM, Olig2 cooperates with pro-tumor factors to sustain tumor cell self-renewal. Limited activation of angiogenic and invasive factors in PA further contributes to its benign nature, as Olig2 does not strongly activate VEGF, MMPs, or EMT genes. In contrast, in GBM, Olig2 induces these genes, enhancing tumor progression. The Olig2-STAT3 interaction in PA remains tumor-suppressive, maintaining astrocyte differentiation, while in GBM, its dysregulation fosters tumorigenic plasticity. Olig2 functions as a context-dependent regulator in gliogenesis. In PA, its activity is tightly regulated by PAX6, STAT3, and NFIA, ensuring differentiation and preventing tumorigenic plasticity. In GBM, loss of these regulatory mechanisms allows Olig2 to sustain tumor cell self-renewal, plasticity, and therapy resistance. The Gliogenic Framework explains why Olig2 remains a differentiation-promoting factor in PA, preventing it from transitioning to a malignant phenotype. Unlike GBM, where Olig2-driven plasticity fuels tumor progression, PA retains gliogenic regulators that suppress Olig2-driven progenitor reprogramming, ensuring it remains benign and slow-growing [82, 83].

### Impact of Gliogenic Regulators on Major Signaling Pathways in Pilocytic Astrocytoma

#### MAPK/ERK Pathway, PI3K/AKT/mTOR Pathway, JAK/STAT Pathway, Shh, Notch

##### 1. MAPK/ERK Pathway

The Mitogen-Activated Protein Kinase (MAPK)/Extracellular Signal-Regulated Kinase (ERK) pathway plays a central role in cell proliferation, differentiation, and survival. In Pilocytic Astrocytoma (PA), the MAPK/ERK pathway is constitutively activated, often due to BRAF fusions (e.g., KIAA1549::BRAF), yet PA remains benign and localized rather than invasive like glioblastoma (GBM). The Gliogenic Framework provides insights into how MAPK/ERK activation in PA is regulated by gliogenic transcription factors, suppressors, and differentiation signals, preventing malignant transformation [84].

In contrast to GBM, where MAPK/ERK activation drives aggressive growth and therapy resistance, PA maintains a controlled balance through differentiation signals. PA is characterized by BRAF fusion-driven MAPK/ERK activation, whereas GBM exhibits EGFR amplification, PTEN loss, and KRAS mutations, which lead to uncontrolled proliferation and invasion. In PA, astrocytic differentiation remains intact, and tumor growth is slow and well-circumscribed, with limited angiogenesis and therapeutic resistance. GBM, on the other hand, is highly invasive, exhibits strong VEGF-driven neovascularization, and resists treatment due to redundancy in signaling pathways [85].

Several gliogenic factors play a role in controlling MAPK/ERK activity in PA. PAX6, a crucial gliogenic differentiation factor, suppresses uncontrolled MAPK/ERK signaling and directs cells toward an astrocytic lineage. Its loss in GBM enables MAPK/ERK-driven dedifferentiation and tumor plasticity. STAT3, which plays a dual role in differentiation and oncogenic plasticity, promotes differentiation in PA by synergizing with MAPK/ERK, whereas in GBM, it sustains tumor stemness and invasion. The NFIA, NFIB, and NFIX transcription factors reinforce astrocytic differentiation in PA by modulating MAPK/ERK activity. In GBM, reduced NFIA/NFIX levels allow MAPK/ERK to drive glioblastoma-initiating cell proliferation. Additionally, HOPX, a differentiation factor and MAPK/ERK suppressor, limits excessive pathway activation in PA, preventing progression to high-grade glioma, while its loss in GBM enables glioma cell self-renewal. MAPK/ERK-associated oncogenic factors are suppressed in PA but active in GBM. SOX9 and SOX10, which are involved in neural stemness and dedifferentiation, are kept in check by PAX6 in PA, preventing a progenitor-like state. In GBM, these factors cooperate with MAPK/ERK to enhance tumor-initiating properties. Notch signaling, which can have both gliogenic and oncogenic roles, favors differentiation in PA but amplifies MAPK/ERK oncogenic signaling in GBM, increasing tumor plasticity. Additionally, VEGF and MMPs, key regulators of angiogenesis and invasion, are not strongly induced by MAPK/ERK in PA, thereby limiting tumor vascularization. In contrast, GBM exploits MAPK/ERK to upregulate VEGF and MMPs, driving aggressive angiogenesis and infiltration. Certain MAPK/ERK-driven pathways, which are oncogenic in GBM, remain restricted in PA. The JAK-STAT3 axis, which fuels tumor progression in GBM, remains transient and non-oncogenic in PA. TGF-β signaling, a key driver of epithelial-to-mesenchymal transition (EMT), migration, and invasion, is not fully activated in PA, whereas in GBM, it promotes tumor cell motility and therapy resistance. Similarly, Wnt/β-catenin signaling, which sustains stemness and therapy resistance in GBM, supports differentiation rather than tumorigenicity in PA [86].

Despite the strong activation of MAPK/ERK, PA does not progress to GBM due to several key factors. Gliogenic transcription factors such as PAX6, NFIA, and STAT3 restrict MAPK/ERK from promoting neural progenitor-like plasticity. Unlike in GBM, where these factors are downregulated, their presence in PA ensures that MAPK/ERK supports differentiation rather than dedifferentiation. Additionally, MAPK/ERK activation in PA is insufficient to drive invasion, as it does not induce VEGF/MMP-mediated tumor spread. In contrast, GBM leverages MAPK/ERK in conjunction with TGF-β and Notch to sustain angiogenesis and tumor infiltration. Therapy resistance is also limited in PA, as it lacks the compensatory pathways that enable GBM cells to evade MAPK inhibitors. Finally, PAX6 and HOPX function as tumor suppressors, preventing MAPK/ERK from driving excessive proliferation in PA, whereas their loss in GBM facilitates an oncogenic state [87].

The Gliogenic Framework provides key insights into why MAPK/ERK activation in PA does not lead to malignant transformation. Gliogenic differentiation factors such as PAX6, NFIA, and STAT3 suppress MAPK/ERK-driven dedifferentiation, ensuring that PA cells retain their astrocytic identity. Unlike GBM, PA does not strongly activate VEGF, MMPs, or EMT factors, limiting its invasive potential. Tumor suppressors such as HOPX and Notch prevent MAPK/ERK from sustaining progenitor-like tumor cells. Additionally, PA lacks the additional mutations seen in GBM, such as EGFR amplification, PTEN loss, and KRAS mutations, which make MAPK/ERK a malignant driver. As a result, MAPK/ERK in PA remains a differentiation-supporting pathway rather than an oncogenic force, ensuring the tumor remains slow-growing, localized, and benign [88, 89].

##### 2. PI3K/AKT/mTOR Pathway

The PI3K/AKT/mTOR signaling pathway is a critical regulator of cell survival, proliferation, metabolism, and differentiation. Dysregulation of this pathway is frequently observed in aggressive gliomas such as glioblastoma (GBM), where it drives stemness, invasion, and therapy resistance. However, in Pilocytic Astrocytoma (PA), although PI3K/AKT/mTOR activation is present, it does not lead to a malignant phenotype. Applying the Gliogenic Framework to PI3K/AKT/mTOR in PA allows us to investigate how gliogenic transcription factors (TFs), differentiation signals, and tumor suppressors modulate this pathway, maintaining PA’s benign and well-circumscribed nature instead of progressing to GBM [90].

Comparing PA and GBM, the activation of the PI3K/AKT/mTOR pathway is moderate and controlled by differentiation factors in PA, whereas in GBM, it is hyperactivated, promoting invasion and therapy resistance. The primary genetic driver in PA is BRAF fusion (KIAA1549::BRAF), while in GBM, it includes EGFR amplification, PTEN loss, and PIK3CA mutations. mTOR signaling in PA supports astrocytic differentiation, whereas in GBM, it drives tumor plasticity and therapy resistance. Angiogenesis is limited in PA due to low VEGF expression, while GBM exhibits high VEGF-driven neovascularization. Cellular invasion remains minimal in PA, keeping the tumor well-circumscribed, while GBM demonstrates high migratory potential. Additionally, therapy resistance is generally low in PA, making it more sensitive to inhibitors, whereas GBM exhibits strong resistance due to compensatory pathways [91].

Gliogenic transcription factors play a crucial role in restricting PI3K/AKT/mTOR activation in PA. PAX6 inhibits excessive PI3K/AKT signaling, preventing dedifferentiation into a progenitor-like state, while its loss in GBM allows PI3K/AKT to sustain stemness and therapy resistance. STAT3 in PA cooperates with PI3K/AKT to promote astrocytic differentiation, but in GBM, its hyperactivation drives PI3K/AKT-mediated tumor progression and invasion. NFIA/NFIX modulate PI3K/AKT to maintain PA’s differentiated astrocytic phenotype, whereas their loss in GBM enables PI3K/AKT to promote stem-like features and therapy resistance. HOPX directly inhibits PI3K hyperactivation, preventing progression to high-grade glioma, but in GBM, HOPX is often downregulated, allowing PI3K/AKT-driven malignancy. PI3K/AKT/mTOR-associated oncogenic factors that are suppressed in PA include SOX9 and SOX10, which, in PA, are inhibited by PAX6 to prevent PI3K/AKT-driven glioma stem-like properties, whereas in GBM, they cooperate with PI3K/AKT to sustain tumor plasticity and invasion. Notch signaling in PA interacts with PI3K to promote astrocytic differentiation, while in GBM, it amplifies PI3K/AKT signaling, enhancing tumor growth and therapy resistance. VEGF and MMPs, which contribute to angiogenesis and invasion, are not strongly induced via PI3K/AKT in PA, limiting tumor vascularization. However, in GBM, PI3K/AKT drives high VEGF/MMP expression, promoting invasive angiogenesis [92].

PI3K/AKT/mTOR-driven pathways that are oncogenic in GBM but restricted in PA include JAK-STAT3, TGF-β/EMT, and Wnt/β-catenin. In PA, PI3K/AKT-driven JAK-STAT3 activation is moderate and differentiation-supporting, whereas in GBM, hyperactive PI3K/AKT-STAT3 signaling sustains glioblastoma stem-like cells. The TGF-β/EMT axis in PA does not fully activate, preventing EMT-driven invasion, while in GBM, PI3K/AKT-TGF-β crosstalk promotes invasion and glioma progression. Wnt/β-catenin interactions in PA favor differentiation, while in GBM, they sustain tumor plasticity and therapy resistance [93].

PI3K/AKT/mTOR-driven PA remains benign instead of progressing to GBM due to several factors. Gliogenic transcription factors such as PAX6, NFIA, and STAT3 restrain PI3K/AKT from driving progenitor-like dedifferentiation, while their loss in GBM enables tumor progression. PI3K/AKT in PA does not strongly activate VEGF/MMP-driven tumor spread, whereas in GBM, it cooperates with TGF-β and Notch to sustain invasion. Limited activation of therapy resistance mechanisms in PA ensures sensitivity to PI3K inhibitors due to the lack of compensatory pathways, unlike GBM, where PI3K/AKT-driven EGFR, Wnt, and JAK-STAT3 cross-activation confers resistance. Additionally, PAX6 and HOPX suppress PI3K/AKT-driven proliferation in PA, ensuring that PI3K/AKT promotes differentiation rather than uncontrolled growth, whereas in GBM, their loss enables oncogenic transformation.

The Gliogenic Framework provides key insights into why PI3K/AKT/mTOR activation in PA does not lead to malignant transformation. Gliogenic differentiation factors such as PAX6, NFIA, and STAT3 suppress PI3K/AKT-driven dedifferentiation. PI3K/AKT in PA does not strongly activate VEGF, MMPs, or EMT factors, limiting invasion. Tumor suppressors such as HOPX and Notch prevent PI3K/AKT from sustaining glioma stem-like cells. Unlike GBM, PA lacks the additional mutations, including EGFR amplification, PTEN loss, and PIK3CA mutations, that make PI3K/AKT oncogenic. Consequently, PI3K/AKT in PA remains a differentiation-supporting pathway rather than a malignant driver, ensuring slow growth, localized behavior, and a benign nature [94, 95].

#### 3. JAK/STAT Pathway

The JAK/STAT pathway is a key regulator of astrocyte differentiation, gliogenesis, immune response, and tumor progression. It plays a dual role in gliomas: in Pilocytic Astrocytoma (PA), JAK/STAT signaling supports astrocytic differentiation and limited proliferation, helping maintain its benign and localized nature, whereas in Glioblastoma (GBM), the same pathway is hyperactivated, driving stemness, tumor plasticity, therapy resistance, and invasiveness. Applying the Gliogenic Framework to JAK/STAT in PA allows us to explore how gliogenic transcription factors (TFs), tumor suppressors, and differentiation signals modulate this pathway, preventing PA from progressing into a high-grade glioma (HGG) like GBM [96].

JAK/STAT activation differs significantly between PA and GBM. In PA, activation remains moderate and differentiation-supportive, with BRAF fusion (KIAA1549::BRAF) as the primary genetic driver. STAT3 promotes astrocytic differentiation, inflammatory cytokines like IL-6 are low, and stemness induction is limited, favoring mature astrocytes. This results in minimal cellular invasion and a well-circumscribed tumor, with sensitivity to inhibitors due to the absence of strong JAK/STAT-PI3K/AKT cross-signaling. In contrast, GBM features hyperactivated JAK/STAT signaling driven by EGFR amplification, PTEN loss, and IL-6 upregulation. STAT3 in GBM drives tumor plasticity and stemness, with chronic inflammation and immune evasion, supporting glioma stem-like cells (GSCs) and enhancing migratory potential. Therapy resistance in GBM is strong due to cross-signaling between JAK/STAT and PI3K/AKT. Several gliogenic transcription factors regulate JAK/STAT in PA, preventing its oncogenic transformation. PAX6 acts as a JAK/STAT inhibitor and differentiation promoter, ensuring differentiation rather than tumor progression. In GBM, PAX6 is often lost, allowing JAK/STAT to drive glioma stem-like properties. NFIA and NFIX enhance STAT3’s differentiation role in PA, suppressing tumor plasticity, whereas their downregulation in GBM permits JAK/STAT to maintain tumor heterogeneity. HOPX functions as a tumor suppressor by restraining STAT3 and modulating JAK/STAT to prevent excessive proliferation in PA; in GBM, its loss enables aggressive tumor growth. Additionally, SOX9 and SOX10, known neural stemness regulators, are suppressed by PAX6 in PA, preventing JAK/STAT from inducing glioma stem-like properties, whereas in GBM, SOX9/10 amplify JAK/STAT-driven tumor plasticity [97, 98].

JAK/STAT-associated oncogenic factors that are suppressed in PA further contribute to its benign nature. IL-6, a key cytokine in GBM progression, remains low in PA, preventing chronic JAK/STAT-driven inflammation. In contrast, IL-6 upregulation in GBM sustains JAK/STAT activation and immune evasion. EGFR, a potent JAK/STAT activator in high-grade gliomas, is limited in PA, whereas its mutations in GBM strongly activate JAK/STAT, fueling malignancy.

Similarly, VEGF and MMPs, critical for angiogenesis and invasion, are not induced via JAK/STAT in PA, keeping the tumor localized, while GBM exploits JAK/STAT to drive high VEGF/MMP expression, promoting invasion.

Pathways driven by JAK/STAT that are oncogenic in GBM remain restricted in PA. The JAK/STAT-PI3K/AKT crosstalk, responsible for therapy resistance and tumor plasticity, is limited in PA, preserving sensitivity to treatment, whereas in GBM, this signaling sustains glioma stem-like cells and therapy resistance. The JAK/STAT-TGF-β interaction, which drives epithelial-to-mesenchymal transition, migration, and invasion, does not fully activate in PA, preventing invasive transformation, but in GBM, it enhances tumor cell invasion and mesenchymal transition. Additionally, Wnt/β-Catenin signaling remains moderate in PA, restricting excessive JAK/STAT-driven stemness, whereas in GBM, Wnt cooperates with JAK/STAT to sustain glioma stem-like cell populations [99].

Several factors explain why JAK/STAT-driven PA remains benign instead of progressing to GBM. Gliogenic transcription factors such as PAX6, NFIA, and STAT3 restrain JAK/STAT from inducing tumor plasticity, whereas their loss in GBM enables JAK/STAT to drive invasion and therapy resistance. In PA, JAK/STAT does not fully activate pro-tumor inflammatory pathways, with low IL-6 levels preventing chronic JAK/STAT-driven immune suppression, while in GBM, high IL-6 creates an immune-suppressive microenvironment. PA also exhibits limited activation of therapy resistance mechanisms, maintaining sensitivity to JAK inhibitors due to restricted PI3K/AKT and Wnt crosstalk, whereas GBM cells resist therapy through JAK/STAT-PI3K/AKT-Wnt synergy. PAX6 and HOPX further suppress JAK/STAT-driven proliferation in PA, ensuring differentiation rather than uncontrolled growth, whereas in GBM, their loss allows JAK/STAT to sustain glioma stemness. The Gliogenic Framework provides key insights into why JAK/STAT activation in PA does not lead to malignant transformation. Gliogenic differentiation factors such as PAX6, NFIA, and STAT3 suppress JAK/STAT-driven tumor plasticity, while JAK/STAT in PA does not strongly activate VEGF, MMPs, or Wnt, limiting invasion. Tumor suppressors like HOPX prevent JAK/STAT from sustaining glioma stem-like cells. Unlike GBM, PA lacks additional oncogenic drivers such as EGFR amplification and IL-6 overexpression that make JAK/STAT oncogenic. As a result, JAK/STAT in PA remains a differentiation-supporting pathway rather than a malignant driver, ensuring slow growth, localized behavior, and a benign nature [100, 101].

##### 4. SHH

The Sonic Hedgehog (Shh) pathway is a key regulator of gliogenesis, neural progenitor cell fate, and astrocyte maturation. While Shh hyperactivation is a driver of high-grade gliomas such as Glioblastoma, Medulloblastoma, and Hedgehog-driven Gliomas, its role in Pilocytic Astrocytoma (PA) remains controlled and differentiation-supportive. This contributes to PA’s benign and localized nature rather than malignant transformation. Applying the Gliogenic Framework to the Shh pathway in PA allows for an investigation into how Shh signaling contributes to the onset and slow-growing nature of PA, why Shh-driven PA remains benign instead of progressing to high-grade gliomas, and which gliogenic transcription factors, tumor suppressors, and differentiation cues regulate Shh in PA to prevent malignancy [102].

In PA, Shh activation is moderate and supports gliogenesis, whereas in Hedgehog-driven gliomas, it is hyperactivated and drives proliferation. The primary genetic driver in PA is the BRAF fusion (KIAA1549::BRAF), while malignant gliomas often exhibit PTCH1 or SMO mutations. Gli1/2 in PA supports astrocyte differentiation, while in malignant gliomas, it drives tumorigenesis and invasiveness. Stemness induction is limited in PA, favoring mature astrocytes, while it is strongly sustained in malignant gliomas, supporting glioma stem-like cells (GSCs). The tumor microenvironment in PA is well-circumscribed with low inflammation, whereas it is pro-tumorigenic and highly inflamed in malignant gliomas. PA remains generally sensitive to inhibitors, while malignant gliomas develop resistance through Shh-PI3K/AKT cross-signaling [103].

PAX6 prevents Shh from inducing excessive proliferation and maintains a differentiation-supportive state, whereas in malignant gliomas, loss of PAX6 leads to unchecked Shh activation, promoting GSCs and tumor plasticity. NFIA and NFIX modulate Shh-activated Gli1/2 to drive astrocyte maturation instead of tumorigenesis. In GBM, their downregulation allows Shh to sustain glioma cell proliferation. HOPX limits Shh-driven stemness and proliferation in PA, keeping the tumor benign, whereas in GBM, HOPX suppression enhances Shh-PI3K/AKT cross-talk, increasing therapy resistance. PAX6 suppresses SOX9/10 in PA, preventing Shh from inducing a stem-like phenotype, whereas in GBM, SOX9/10 amplify Shh signaling, sustaining tumor cell plasticity. Oncogenic factors associated with Shh activation are suppressed in PA. Gli1/2 activation remains moderate in PA, ensuring astrocytic differentiation rather than proliferation, whereas in GBM, their hyperactivation drives tumorigenesis, invasion, and therapy resistance. PTCH1 or SMO mutations, which are absent in PA, contribute to full-scale Shh-driven oncogenesis in Medulloblastoma and Shh-driven Gliomas. PI3K/AKT activation via Shh remains limited in PA, maintaining treatment sensitivity, while in GBM, Shh-PI3K/AKT signaling sustains GSCs and therapy resistance [104].

Certain Shh-driven pathways that contribute to oncogenesis in GBM are restricted in PA. The Shh-PI3K/AKT interaction is weak in PA, limiting cell proliferation, whereas in GBM, this cooperation supports tumor stemness and resistance to inhibitors. Shh-TGF-β crosstalk, which contributes to epithelial-mesenchymal transition (EMT), migration, and invasion, does not fully activate in PA, preventing invasive transformation. However, in GBM, this crosstalk enables tumor cell invasion and mesenchymal transition. Shh-Wnt/β-Catenin crosstalk remains moderate in PA, preventing excessive Shh-driven stemness, whereas in GBM, Wnt cooperates with Shh to sustain GSC populations. The benign nature of Shh-driven PA can be attributed to several factors. Gliogenic transcription factors such as PAX6, NFIA, and STAT3 prevent Shh from inducing tumor plasticity, whereas their loss in GBM enables Shh to drive invasion and therapy resistance. Limited Gli1/2 activity in PA prevents high proliferation, whereas in GBM, these factors drive oncogenic transformation. Therapy resistance mechanisms remain limited in PA, ensuring sensitivity to Shh inhibitors, while in GBM, Shh-PI3K/AKT-Wnt synergy facilitates resistance. PAX6 and HOPX in PA suppress Shh-driven proliferation, ensuring differentiation rather than uncontrolled growth, whereas in GBM, loss of these factors enables Shh to sustain glioma stemness [105].

The Gliogenic Framework provides key insights into why Shh activation in PA does not lead to malignant transformation. Gliogenic differentiation factors such as PAX6, NFIA, and STAT3 suppress Shh-driven tumor plasticity. In PA, Shh does not strongly activate TGF-β, VEGF, or Wnt, limiting invasion. Tumor suppressors like HOPX prevent Shh from sustaining glioma stem-like cells. Unlike GBM, PA lacks additional oncogenic drivers such as PTCH1/SMO mutations and IL-6 overexpression that make Shh oncogenic. Thus, Shh in PA remains a differentiation-supporting pathway rather than a malignant driver, ensuring slow growth, localized behavior, and a benign nature [106, 107].

##### 5. Notch

The Notch signaling pathway is a crucial regulator of gliogenesis, neural progenitor maintenance, and astrocytic differentiation. In high-grade gliomas such as glioblastoma and medulloblastoma, Notch hyperactivation contributes to glioma stem cell (GSC) maintenance, invasion, and therapy resistance. However, in Pilocytic Astrocytoma (PA), Notch signaling remains moderate and differentiation-supportive, playing a key role in maintaining its benign and localized nature rather than driving aggressive growth. Applying the Gliogenic Framework to the Notch pathway in PA allows us to investigate how Notch signaling contributes to PA onset and slow growth, why Notch-driven PA remains benign instead of progressing to high-grade gliomas, and which gliogenic transcription factors (TFs) and tumor suppressors regulate Notch in PA to prevent malignancy [108].

In PA, Notch activation is moderate and supports differentiation, while in high-grade gliomas, it is hyperactivated and drives tumorigenesis. The primary genetic driver in PA is the BRAF fusion (KIAA1549::BRAF), whereas in malignant gliomas, NOTCH1/NOTCH2 amplifications and DLL3 overexpression are common. Gliogenic TFs such as PAX6, NFIA, and STAT3 restrict Notch-induced tumor plasticity in PA, but their loss in high-grade gliomas allows Notch to sustain glioma stemness. Notch-JAK/STAT crosstalk in PA is limited, preventing Notch-induced immune suppression, whereas in malignant gliomas, it is hyperactive and supports tumor immune evasion. Similarly, Notch-PI3K/AKT crosstalk is restricted in PA, limiting proliferation, while in high-grade gliomas, it enhances glioma survival and therapy resistance. Stemness induction in PA is limited and favors mature astrocytes, whereas in gliomas, it sustains glioma stem-like cells (GSCs). The tumor microenvironment (TME) in PA is non-invasive with a low inflammatory response, in contrast to the pro-tumorigenic and highly inflammatory TME in malignant gliomas. PA remains generally sensitive to Notch inhibitors, whereas high-grade gliomas develop resistance through Notch-PI3K/AKT cross-signaling [109, 110].

PAX6 suppresses Notch-driven progenitor expansion and enhances differentiation in PA, whereas its loss in GBM allows Notch to sustain glioma stem cell-like states, increasing therapy resistance. NFIA and NFIX prevent Notch from sustaining neural progenitors in PA, directing cells toward astrocytic differentiation, whereas their loss in GBM enables Notch to drive tumor plasticity. HOPX limits Notch-driven stemness and tumor cell renewal in PA, but in GBM, its suppression enhances Notch-PI3K/AKT crosstalk, supporting glioma stem-like cells (GSCs). STAT3 in PA restricts

Notch activity to maintain a differentiation-supportive role, whereas in GBM, hyperactivated STAT3-Notch signaling sustains tumor survival and immune evasion [111].

NOTCH1/NOTCH2 activation in PA remains moderate, ensuring differentiation rather than excessive growth, whereas their amplification in GBM drives tumor invasion, GSC maintenance, and therapy resistance. HES1 and HEY1 in PA function in a restricted manner to support astrocyte maturation, whereas in GBM, their uncontrolled activation sustains glioma stem-like properties. Notch-PI3K/AKT crosstalk in PA is weak, preventing excessive proliferation, whereas in GBM, this interaction supports glioma stem-like cells and resistance to inhibitors. DLL3 overexpression is absent in PA, preventing Notch-induced tumorigenesis, but in GBM, DLL3 sustains Notch signaling and tumor survival. Notch-driven pathways that are oncogenic in GBM remain restricted in PA. Notch-PI3K/AKT crosstalk is weak in PA, limiting cell proliferation, whereas in GBM, it sustains glioma stem-like cells and therapy resistance. Notch-Wnt/β-catenin crosstalk remains moderate in PA, preventing excessive Notch-driven stemness, whereas in GBM, Wnt cooperates with Notch to sustain glioma stem-like cell populations. Notch-TGF-β crosstalk does not fully activate in PA, preventing invasive transformation, whereas in GBM, it enables tumor cell invasion and mesenchymal transition. PA remains benign instead of progressing to GBM due to several factors. Gliogenic transcription factors such as PAX6, NFIA, and STAT3 prevent Notch from inducing tumor plasticity, whereas their loss in GBM enables Notch to drive invasion and therapy resistance. Notch in PA does not fully activate pro-tumor pathways, as limited Notch1/2 activity prevents high proliferation, whereas in GBM, Notch hyperactivation drives oncogenic transformation. PA has limited activation of therapy resistance mechanisms, remaining sensitive to Notch inhibitors due to restricted PI3K/AKT and Wnt crosstalk, whereas GBM cells resist therapy through Notch-PI3K/AKT-Wnt synergy. PAX6 and HOPX in PA suppress Notch-driven proliferation, ensuring that Notch supports differentiation rather than uncontrolled growth, whereas in GBM, loss of PAX6/HOPX enables Notch to sustain glioma stemness. The Gliogenic Framework provides key insights into why Notch activation in PA does not lead to malignant transformation. Gliogenic differentiation factors such as PAX6, NFIA, and STAT3 suppress Notch-driven tumor plasticity. Notch in PA does not strongly activate TGF-β, VEGF, or Wnt, limiting invasion. Tumor suppressors such as HOPX prevent Notch from sustaining glioma stem-like cells. By maintaining a differentiation-supportive role and restricting oncogenic crosstalk, Notch signaling in PA remains a regulator of astrocytic maturation rather than a driver of aggressive tumor growth [112, 113].

## Discussion

### PA Oncogenesis and the Proliferation-Differentiation Balance in Glial Cells

Pilocytic Astrocytoma (PA) oncogenesis disrupts the proliferation-differentiation balance in glial cells, primarily through dysregulated MAPK/ERK signaling. Normally, gliogenic pathways such as BMP, PAX6, STAT3, and NFIA regulate glial cell differentiation, ensuring astrocytes maintain their supportive and homeostatic roles. However, in PA, constitutive activation of MAPK/ERK, due to BRAF fusions or NF1 loss, skews this balance. This leads to uncontrolled proliferation, as MAPK/ERK hyperactivation promotes sustained cell division, preventing glial cells from reaching full differentiation. The loss of tumor suppressors such as NF1, PTEN, and TP53 removes negative regulation of cell cycle progression, while upregulation of PI3K/AKT/mTOR and JAK/STAT, though limited, supports metabolic changes that favor proliferation over differentiation.

At the same time, differentiation is compromised due to reduced PAX6 and NFIA function, impairing differentiation signals and preventing glial cells from exiting the cell cycle. SOX10 and STAT3, though gliogenic, exhibit oncogenic behaviors in PA, sustaining astrocyte-like but undifferentiated states. Additionally, HOPX downregulation prevents terminal differentiation, further disrupting astrocytic fate commitment. Despite these oncogenic disruptions, PA remains benign because its cells do not fully revert to a stem-like state due to preserved gliogenic programs. Unlike high-grade gliomas, PA lacks aggressive angiogenic drivers and does not breach basement membranes, maintaining low invasiveness. Gliogenic players such as PAX6, NFIA, and BMP continue to function, preventing excessive stemness and allowing partial differentiation.

PA oncogenesis favors proliferation over differentiation but remains partially gliogenic. While glial cells fail to mature fully, their tumorigenic transformation is limited by residual differentiation signals. This incomplete transformation prevents PA from acquiring high-grade glioma characteristics, ensuring its benign, non-invasive nature.

#### Key Findings Emerging from This Study

1. Gliogenic Framework as a Tumor-Modulating Network

- The study highlights how gliogenic transcription factors (e.g., PAX6, SOX10, NFIs) and signaling pathways (Notch, Shh, JAK/STAT) not only drive glial differentiation but also act as modulators that suppress malignancy in PA.
2. Dual Nature of Oncogenic Pathways in PA

- Unlike other gliomas, PA exhibits oncogenic activation (e.g., MAPK/ERK, PI3K/AKT/mTOR, JAK/STAT) while retaining differentiation signals, preventing unchecked proliferation and dedifferentiation into high-grade malignancies.
3. PTEN and TP53 Function as Stability Factors

- PA maintains a non-malignant state due to the relative stability of PTEN and TP53, which counterbalance the proliferative effects of oncogenic pathways. Their partial loss or mutation does not cause immediate transformation, unlike in glioblastoma.
4. BRAF Mutation and Controlled Oncogenesis

- The BRAF fusion (KIAA1549::BRAF) drives proliferation without disrupting differentiation programs, restricting aggressive invasion—a phenomenon unique to PA compared to other BRAF-driven cancers.
5. Notch and Shh in Tumor Maintenance

- Notch and Shh, while oncogenic in high-grade gliomas, exhibit a pro-differentiation role in PA, reinforcing the benign nature by sustaining glial cell identity.
6. Limited Angiogenic and Invasive Potential

- PA lacks the extensive vascular proliferation and extracellular matrix remodeling seen in high-grade astrocytomas, reducing its metastatic capacity.
7. Stemness Factors in PA vs. GBM

- Unlike glioblastoma, PA does not heavily rely on SOX2, OCT4, or Nanog, explaining why it retains glial differentiation cues and does not dedifferentiate into a more aggressive phenotype.

These findings deepen our understanding of PA’s unique proliferation-differentiation balance, distinguishing it from malignant gliomas.

### Glial Cells’ Predisposition to Pilocytic Astrocytoma (PA) Risk

Glial cells, particularly astrocyte precursors and neural progenitor cells (NPCs), exhibit a predisposition to Pilocytic Astrocytoma (PA) development due to their inherent proliferative and differentiation capacities. Several factors contribute to this susceptibility:

1. Neural Progenitor Vulnerability to Oncogenic Activation

- PA predominantly arises in pediatric populations, where neural progenitor cells (NPCs) are actively proliferating. These cells have a high sensitivity to oncogenic mutations (e.g., BRAF fusion, NF1 loss, FGFR1 activation) that drive MAPK/ERK hyperactivation, initiating PA formation.
2. MAPK/ERK Pathway as a Primary Driver

- The strong dependence of glial progenitors on MAPK/ERK signaling for differentiation and proliferation makes them prone to PA initiation when BRAF fusions or NF1 loss occur. However, unlike glioblastoma, PA does not undergo full dedifferentiation, allowing it to remain benign.
3. Differentiation-Linked Oncogenesis

- Unlike high-grade gliomas, PA originates in cells that still retain glial differentiation programs. SOX10, NFIs, and PAX6 play key roles in guiding astrocytic lineage commitment, preventing aggressive dedifferentiation and malignant transformation.
4. Role of Tumor-Suppressor Stability (PTEN, TP53)

- While PA shows some oncogenic alterations, PTEN and TP53 remain relatively intact, preventing unrestricted proliferation and promoting cell-cycle exit upon differentiation, reinforcing the benign phenotype.
5. Restricted Angiogenesis and Invasion

- Unlike high-grade gliomas, PA does not exhibit extensive angiogenesis or extracellular matrix remodeling, limiting its invasive potential. This stems from glial precursors’ intrinsic non-invasive properties.

Thus, glial cell lineage-specific vulnerabilities, combined with the selective activation of MAPK/ERK-driven proliferation without full oncogenic dedifferentiation, make PA a distinct, benign glioma.

### Clinical Implications of the Findings of This Study

1. Targeted Therapeutics for PA

- Understanding the role of MAPK/ERK hyperactivation (BRAF fusion, NF1 loss) suggests that MEK inhibitors could serve as effective therapeutic strategies for PA, minimizing unnecessary chemotherapy or radiation.
2. Biomarker-Driven Diagnosis

- The identification of glial-specific transcription factors (SOX10, PAX6, NFIs) and signaling pathways (Notch, Shh, JAK/STAT) as regulators of PA progression provides biomarkers for early detection and prognosis.
3. Preventing Malignant Transformation

- Despite its benign nature, PA has rare cases of malignant progression. Insights into PTEN/TP53 stability and angiogenic restrictions may help identify patients at risk for transformation and guide preventive interventions.
4. Refining Surgical and Post-Surgical Strategies

- Understanding PA’s limited invasiveness supports the current gross-total resection approach, while future research could explore post-surgical molecular monitoring to predict recurrence risks.

### Implications in PA Oncogenesis Research

1. New Perspective on Gliogenic Oncogenesis

- This study highlights that PA emerges from differentiation-linked oncogenesis, in contrast to other gliomas, which undergo dedifferentiation-driven malignancy.
2. MAPK-Driven Benign Oncogenesis

- Unlike glioblastoma, PA sustains proliferation without full transformation, reinforcing that oncogenesis does not always lead to malignancy and prompting deeper research into controlled oncogenic pathways.
3. Role of Tumor Microenvironment in PA Benignity

- The findings suggest that PA’s non-invasive nature is linked to glial-intrinsic differentiation programs, offering a new dimension to glioma microenvironment research.
4. Potential for Differentiation-Based Therapies

- Since PA retains glial differentiation markers, differentiation-inducing therapies (e.g., modulating SOX10, NFIs, PAX6) could be explored as a treatment strategy to further reduce proliferation.

These findings significantly advance the current understanding of PA and its unique oncogenesis, paving the way for precision medicine and future glioma research.

## Conclusion

The gliogenic framework provides critical insights into how transcription factors, signaling cascades, and cellular mechanisms regulate PA’s onset, differentiation, and non-malignant behavior. This framework defines the pathways involved in glial differentiation, progenitor cell maintenance, and tumorigenesis, where these players interact in a way that initiates tumor formation without allowing malignant progression. PA development is primarily driven by aberrant MAPK/ERK activation, usually through the KIAA1549::BRAF fusion. However, unlike malignant gliomas, PA does not acquire additional oncogenic mutations such as TERT promoter mutations, CDKN2A/B deletions, or TP53 loss, preventing aggressive growth. Gliogenic transcription factors such as PAX6, NFIA, NFIB, and STAT3 promote astrocytic differentiation, ensuring that PA cells do not dedifferentiate into stem-like, invasive glioma cells. Olig2 and AP-1 remain in a gliogenic, rather than neurogenic, state, reinforcing a controlled glial lineage commitment, while SOX10 does not drive stemness as it does in high-grade gliomas, allowing cells to maintain their mature phenotype. Furthermore, PI3K/AKT/mTOR, JAK/STAT, and Notch signaling remain in a state that supports differentiation rather than proliferation and therapy resistance, while Shh signaling, although active in early gliogenesis, does not reach the levels required for malignant transformation. HOPX suppresses Notch-driven glioma stemness, ensuring PA remains non-aggressive.

Several mechanisms prevent PA from becoming malignant, primarily through the absence of invasive mechanisms, a tumor microenvironment that promotes differentiation and immune surveillance, and genomic stability that prevents malignant evolution. Low TGF-β and MMP activity prevent epithelial-to-mesenchymal transition (EMT) and matrix degradation, restricting invasion. Unlike GBM, PA does not induce VEGF-driven angiogenesis, limiting vascularization and rapid expansion. PA also lacks the immunosuppressive microenvironment seen in malignant gliomas, allowing immune cells to control tumor growth. Reactive astrocytes dominate the tumor microenvironment rather than glioma-associated macrophages (GAMs), preventing a pro-tumor inflammatory niche. Additionally, PA does not accumulate the multiple mutations required for progression to high-grade gliomas, and its low mutation burden ensures that its oncogenic drivers, such as the BRAF fusion, do not trigger further malignant transformation.

The gliogenic framework explains PA’s unique biology: while it originates from gliogenic progenitor cells and activates certain oncogenic pathways, it maintains strong differentiation signals, avoids tumor plasticity, and lacks key genetic alterations that drive high-grade gliomas. Controlled MAPK/ERK activation ensures growth but not malignancy, while gliogenic transcription factors such as PAX6, NFIA, STAT3, and SOX10 support differentiation, preventing dedifferentiation into an aggressive state. Minimal activation of PI3K/AKT, JAK/STAT, Notch, and Shh keeps proliferation and invasion in check, and limited extracellular matrix remodeling, low angiogenesis, and a non-immunosuppressive tumor microenvironment restrict PA’s ability to become invasive. Additionally, genomic stability ensures PA does not acquire secondary mutations that lead to malignancy.

Pilocytic Astrocytoma represents a unique glioma subtype where oncogenic activation is tightly regulated within the gliogenic framework, preventing its progression to a high-grade malignancy. Its developmental trajectory is driven by gliogenic transcription factors and signaling pathways that sustain differentiation, limit invasion, and maintain a stable genome, ensuring its benign nature.

Pilocytic Astrocytoma (PA) is driven by MAPK/ERK activation, yet remains benign due to regulated gliogenic pathways (PAX6, STAT3, SOX10, NFIA). It lacks genetic instability, preventing malignant transformation. Limited PI3K/AKT, JAK/STAT, and Notch signaling restricts invasiveness. Differentiation-promoting factors (PAX6, NFIA) counteract oncogenesis. PA’s non-immunosuppressive microenvironment, low angiogenesis, and restricted proliferation further prevent progression. The absence of stem-like, aggressive properties distinguishes PA from high-grade gliomas, ensuring controlled tumor growth and cellular stability, solidifying its benign nature and non-malignant behavior.

### Why PA Remains Benign?

Pilocytic Astrocytoma (PA) is a slow-growing, well-circumscribed, and generally non-invasive brain tumor that arises primarily in children and young adults. Unlike high-grade gliomas, such as glioblastoma (GBM), PA rarely undergoes malignant transformation, exhibits minimal infiltration, and is often treatable with surgical resection alone.

Unlike aggressive gliomas, PA exhibits moderate activation of key oncogenic signaling pathways without triggering uncontrolled proliferation or tumor stemness. The primary genetic driver of PA is the KIAA1549::BRAF fusion, leading to constitutive activation of the MAPK/ERK pathway. However, this activation remains restricted, sustaining slow growth rather than uncontrolled proliferation. In contrast, GBM and diffuse astrocytomas exhibit multiple oncogenic mutations (e.g., IDH1/2, EGFR, PTEN loss, TERT promoter mutations), driving rapid tumor expansion. While Notch1/2 signaling is active, it remains in a differentiation-supportive state, promoting astrocyte-like features rather than glioma stem cell (GSC) maintenance. PA does not exhibit PTEN loss, preventing excessive PI3K/AKT-driven growth. Additionally, STAT3 activation is controlled, preventing chronic inflammation and immune evasion, which are hallmarks of GBM progression.

Strong differentiation signals in PA restrict tumor plasticity. PAX6, NFIA, and STAT3 ensure that PA cells exit the progenitor state and differentiate into astrocyte-like, non-invasive cells. In contrast, GBM downregulates PAX6 and NFIA, leading to tumor plasticity, self-renewal, and therapy resistance. HOPX suppresses Notch-driven tumor plasticity, preventing a shift toward a glioblastoma-like phenotype. Loss of HOPX in GBM promotes Notch-PI3K/AKT signaling, enabling invasion and therapy resistance.

PA exhibits low invasiveness due to minimal extracellular matrix (ECM) remodeling and suppression of epithelial-to-mesenchymal transition (EMT). Invasive gliomas hijack the TGF-β pathway to promote EMT and matrix metalloproteinase (MMP) activity, enabling infiltration into healthy brain tissue. PA does not fully activate TGF-β, avoiding mesenchymal transition and remaining well-circumscribed. Unlike GBM tumors, which induce VEGF overexpression to drive aberrant vascularization and rapid growth, PA does not exhibit significant VEGF upregulation, ensuring low angiogenesis and controlled proliferation.

The tumor microenvironment (TME) of PA is more favorable compared to GBM. GBM manipulates the TME via immunosuppressive cytokines (IL-10, TGF-β) to evade immune detection, whereas PA does not generate a strongly immunosuppressive TME, allowing the immune system to contain tumor growth. Additionally, the PA microenvironment consists of reactive astrocytes rather than invasive glioma-associated macrophages (GAMs). In contrast, GBM recruits microglia and GAMs to create a pro-tumor niche.

PA also demonstrates genomic stability and a low mutation burden, preventing malignant progression. It lacks the high mutational burden seen in aggressive gliomas. Key oncogenic events in GBM, such as TERT promoter mutations, EGFR amplifications, and PTEN loss, are absent in PA. Without these secondary mutations, PA does not evolve into a higher-grade malignancy.

PA remains benign because its MAPK/ERK activation (BRAF fusion-driven) sustains slow growth rather than aggressive proliferation. Gliogenic differentiation factors (PAX6, NFIA, HOPX, STAT3) restrict Notch and JAK/STAT signaling, preventing glioblastoma-like tumor plasticity. Minimal ECM remodeling and weak TGF-β activity prevent tumor invasion. A favorable immune environment allows natural immune control over tumor growth. Low mutation burden and genomic stability ensure PA does not acquire malignant features. Ultimately, PA maintains a controlled activation of oncogenic pathways (MAPK, Notch, PI3K) while preserving differentiation signals (PAX6, NFIA, STAT3) and avoiding key hallmarks of malignancy, such as invasion, immune suppression, and high mutation burden.

### What Prevents Pilocytic Astrocytoma (PA) from Becoming Malignant?

Pilocytic Astrocytoma (PA) is a low-grade, well-circumscribed, and typically non-invasive glioma that rarely progresses to a malignant form. Unlike high-grade gliomas, such as glioblastoma (GBM), PA exhibits biological, molecular, and microenvironmental features that restrict malignancy. From our analysis of gliogenic frameworks, key signaling pathways, and tumor biology, several factors prevent PA from undergoing malignant transformation.

A key factor is restricted proliferation due to controlled oncogenic pathway activation. The major driver of PA is the KIAA1549::BRAF fusion, which constitutively activates the MAPK/ERK pathway. However, this activation does not reach the threshold needed for high-grade gliomagenesis due to the absence of cooperating mutations such as CDKN2A/B loss or TERT promoter activation. Unlike GBM, which exhibits multiple driver mutations, PA remains genetically stable. Additionally, PA lacks PTEN loss, which keeps PI3K/AKT activation in check. Invasive gliomas rely on hyperactive PI3K/AKT/mTOR signaling to promote growth and survival, whereas PA does not exhibit such chronic activation. Similarly, JAK/STAT signaling is not persistently active, preventing immune evasion and inflammation-induced malignancy. Furthermore, Notch signaling in PA promotes glial differentiation rather than tumor stemness, contrasting with GBM, which hijacks Notch to maintain a therapy-resistant glioma stem cell (GSC) population.

Another critical barrier to malignancy in PA is the presence of strong differentiation signals that suppress tumor plasticity. PAX6, NFIA, and STAT3 promote terminal differentiation of PA cells, preventing dedifferentiation into stem-like, invasive glioma cells. In contrast, GBM loses these differentiation factors, allowing tumor cells to acquire a more plastic, therapy-resistant phenotype. HOPX, another critical factor, inhibits Notch-driven glioma stemness and suppresses the transition to a high-grade glioma state. PA retains HOPX expression, reinforcing its benign nature.

The tumor’s limited invasive potential further ensures its non-malignant behavior. GBM tumors exploit TGF-β signaling and matrix metalloproteinases (MMPs) to invade brain tissue. However, PA does not exhibit significant TGF-β-driven epithelial-to-mesenchymal transition (EMT) or extracellular matrix (ECM) degradation, ensuring that it remains well-circumscribed rather than diffusely infiltrating adjacent brain regions. Additionally, low angiogenesis prevents aggressive growth, as GBM induces VEGF-driven neovascularization, leading to rapid expansion, whereas PA does not show extensive VEGF upregulation, restricting its ability to form an abnormal vascular network.

The tumor microenvironment (TME) also plays a vital role in preventing malignant progression. Unlike GBM, which manipulates the immune system via IL-10, TGF-β, and PD-L1 expression to create an immunosuppressive niche, PA does not suppress the immune response, allowing immune cells to contain tumor growth. Furthermore, while GBM recruits glioma-associated macrophages (GAMs) to promote immune evasion and invasion, PA’s microenvironment consists mostly of reactive astrocytes, which do not actively drive tumor progression. Finally, genomic stability prevents PA from evolving into a more aggressive tumor. High-grade gliomas accumulate multiple genetic alterations, including EGFR amplification, CDKN2A/B deletion, TP53 and PTEN mutations, and TERT promoter activation. PA lacks these secondary mutations, preventing its progression to a malignant state.

Several biological and molecular barriers contribute to preventing PA from becoming malignant. Controlled MAPK/ERK activation due to BRAF fusion prevents rapid proliferation, while differentiation factors such as PAX6, NFIA, STAT3, and HOPX suppress tumor plasticity. Weak TGF-β signaling, low MMP activity, and intact ECM prevent invasion. The tumor’s non-immunosuppressive microenvironment limits aggressive tumor progression, and genomic stability, characterized by a lack of key oncogenic mutations, prevents malignant evolution. Ultimately, PA remains benign because it activates oncogenic pathways in a controlled manner while maintaining strong differentiation cues, avoiding immune suppression, and lacking key drivers of malignancy, such as invasion, angiogenesis, and a high mutation burden.

## Abbreviations

PA: Pilocytic Astrocytoma
GF: Gliogenic Framework
NSCs: Neural Stem Cells
GSCs: Glioma Stem Cells
BRAF: B-Raf Proto-Oncogene, Serine/Threonine Kinase
NF1: Neurofibromin 1
FGFR1: Fibroblast Growth Factor Receptor 1
PTEN: Phosphatase and Tensin Homolog
TP53: Tumor Protein P53
SOX10: SRY-Box Transcription Factor 10
NFI: Nuclear Factor I Family (NFIA, NFIB, NFIX)
STAT3: Signal Transducer and Activator of Transcription 3
PAX6: Paired Box 6
AP-1: Activator Protein 1
Olig2: Oligodendrocyte Transcription Factor 2
MAPK/ERK: Mitogen-Activated Protein Kinase / Extracellular Signal-Regulated Kinase Pathway
PI3K/AKT/mTOR: Phosphoinositide 3-Kinase / Protein Kinase B / Mammalian Target of Rapamycin Pathway
JAK/STAT: Janus Kinase / Signal Transducer and Activator of Transcription Pathway
Shh: Sonic Hedgehog Pathway
TFs: Transcription Factors

## Declarations

### Ethics declarations

#### Ethics approval and consent to participate

Not applicable.

### Consent for publication

Not applicable.

### Data Availability statement

All data generated or analyzed during this study are included in this article.

## Acknowledgements

“Not applicable”.

## Authors’ Information

1. **Ovais Shafi (OS)\*** is the author of the study and was involved in the idea, concept, design, and methodology of the study, literature search and references. He did the writing, editing, and revision of the manuscript. He was involved in drawing the findings, results, conclusions, implications of the study, interpretation of the data and was involved in all aspects of the study. He prepared and wrote discussion, results, conclusions and all areas of the study. OS extracted and analyzed the data. He was involved in critical evaluation, audit of every aspect of the study, data extraction, adherence of the study to relevant PRISMA guidelines, limitations of the study, references, and all others. He was involved in drawing PRISMA Flow Diagram. The author read and approved the manuscript.

2. **Ovais Shafi (OS)\***, MBBS - Sindh Medical College - Dow University of Health Sciences, Karachi, Pakistan. He aspires to become an eminent ‘Physician Scientist’. He is devoted to the research in disease development mechanisms, disease origins and therapeutics. OS is also passionate about multiple research areas including clinical trials, clinical medicine, therapeutics, regenerative medicine, precision medicine including gene therapies, finding disease specific targets for gene therapy, role of disease genomics and epigenetics in diagnosis, management, and therapeutics development. He is dedicated to the field of research and clinical medicine.

Corresponding author: OS

Correspondence to <u>Ovais Shafi</u>

**Raveena (RA)** is also the author of the study and contributed to the writing, editing and revision. She also contributed to the results and conclusions sections of the study along with working on the findings, interpretation of the data and references.

Raveena, MBBS - Sindh Medical College – Jinnah Sindh Medical University, Karachi, Pakistan. She is passionate about research in surgery and disease development mechanisms including neurodegenerative diseases, oncogenesis and others. She is ECFMG Certified. She is passionate about residency in Internal Medicine/Surgery. Her goal is to make significant impact in the field of Research.

## References

1. Bornhorst M, Frappaz D, Packer RJ. Pilocytic astrocytomas. Handb Clin Neurol. 2016;134:329–44. doi: 10.1016/B978-0-12-802997-8.00020-7. PMID: 26948364.

2. Pizzimenti C, Fiorentino V, Germanò A, Martini M, Ieni A, Tuccari G. Pilocytic astrocytoma: The paradigmatic entity in low-grade gliomas (Review). Oncol Lett. 2024 Feb 8;27(4):146. doi: 10.3892/ol.2024.14279. PMID: 38385109; PMCID: PMC10879958.

3. Bilginer B, Narin F, Oguz KK, Uzun S, Soylemezoglu F, Akalan N. Benign cerebellar pilocytic astrocytomas in children. Turk Neurosurg. 2011 Jan;21(1):22–6. PMID: 21294087.

4. Collins VP, Jones DT, Giannini C. Pilocytic astrocytoma: pathology, molecular mechanisms and markers. Acta Neuropathol. 2015 Jun;129(6):775–88. doi: 10.1007/s00401-015-1410-7. Epub 2015 Mar 20. PMID: 25792358; PMCID: PMC4436848.

5. Cler SJ, Skidmore A, Yahanda AT, Mackey K, Rubin JB, Cluster A, Perkins S, Gauvain K, King AA, Limbrick DD, McEvoy S, Park TS, Smyth MD, Mian AY, Chicoine MR, Dahiya S, Strahle JM. Genetic and histopathological associations with outcome in pediatric pilocytic astrocytoma. J Neurosurg Pediatr. 2022 Feb 11;29(5):504–512. doi: 10.3171/2021.9.PEDS21405. PMID: 35148515.

6. Riemenschneider MJ, Reifenberger G. Astrocytic tumors. Recent Results Cancer Res. 2009;171:3–24. doi: 10.1007/978-3-540-31206-2_1. PMID: 19322535.

7. Kessler T, Ito J, Wick W, Wick A. Conventional and emerging treatments of astrocytomas and oligodendrogliomas. J Neurooncol. 2023 May;162(3):471–478. doi: 10.1007/s11060-022-04216-z. Epub 2022 Dec 25. PMID: 36566461; PMCID: PMC10226903.

8. Van de Kelft E. Molecular pathogenesis of astrocytoma and glioblastoma multiforme. Acta Neurochir (Wien). 1997;139(7):589–99. doi: 10.1007/BF01411992. PMID: 9265950.

9. Konopka G, Bonni A. Signaling pathways regulating gliomagenesis. Curr Mol Med. 2003 Feb;3(1):73–84. doi: 10.2174/1566524033361609. PMID: 12558076.

10. Caccese M, Padovan M, D’Avella D, Chioffi F, Gardiman MP, Berti F, Busato F, Bellu L, Bergo E, Zoccarato M, Fassan M, Zagonel V, Lombardi G. Anaplastic Astrocytoma: State of the art and future directions. Crit Rev Oncol Hematol. 2020 Sep;153:103062. doi: 10.1016/j.critrevonc.2020.103062. Epub 2020 Jul 17. PMID: 32717623.

11. Butowski NA, Sneed PK, Chang SM. Diagnosis and treatment of recurrent high-grade astrocytoma. J Clin Oncol. 2006 Mar 10;24(8):1273–80. doi: 10.1200/JCO.2005.04.7522. PMID: 16525182.

12. Furnari FB, Fenton T, Bachoo RM, Mukasa A, Stommel JM, Stegh A, Hahn WC, Ligon KL, Louis DN, Brennan C, Chin L, DePinho RA, Cavenee WK. Malignant astrocytic glioma: genetics, biology, and paths to treatment. Genes Dev. 2007 Nov 1;21(21):2683–710. doi: 10.1101/gad.1596707. PMID: 17974913.

13. Wang Y, Xing H, Guo X, Chen W, Wang Y, Liang T, Wang H, Li Y, Jin S, Shi Y, Liu D, Yang T, Xia Y, Li J, Wu J, Liu Q, Qu T, Guo S, Li H, Zhang K, Wang Y, Ma W. Clinical features, MRI, molecular alternations, and prognosis of astrocytoma based on WHO 2021 classification of central nervous system tumors: A single-center retrospective study. Cancer Med. 2024 Jul;13(13):e7369. doi: 10.1002/cam4.7369. PMID: 38970209; PMCID: PMC11226410.

14. Nahar Metu CL, Sutihar SK, Sohel M, Zohora F, Hasan A, Miah MT, Rani Kar T, Hossain MA, Rahman MH. Unraveling the signaling mechanism behind astrocytoma and possible therapeutics strategies: A comprehensive review. Cancer Rep (Hoboken). 2023 Oct;6(10):e1889. doi: 10.1002/cnr2.1889. Epub 2023 Sep 7. PMID: 37675821; PMCID: PMC10598261.

15. Shafi O, Siddiqui G. Tracing the origins of glioblastoma by investigating the role of gliogenic and related neurogenic genes/signaling pathways in GBM development: a systematic review. World J Surg Oncol. 2022 May 10;20(1):146. doi: 10.1186/s12957-022-02602-5. PMID: 35538578; PMCID: PMC9087910.

16. Sojka C, Sloan SA. Gliomas: a reflection of temporal gliogenic principles. Commun Biol. 2024 Feb 6;7(1):156. doi: 10.1038/s42003-024-05833-2. PMID: 38321118; PMCID: PMC10847444.

17. Nakada M, Kita D, Watanabe T, Hayashi Y, Teng L, Pyko IV, Hamada J. Aberrant signaling pathways in glioma. Cancers (Basel). 2011 Aug 10;3(3):3242–78. doi: 10.3390/cancers3033242. PMID: 24212955; PMCID: PMC3759196.

18. Lin A, Rodriguez FJ, Karajannis MA, Williams SC, Legault G, Zagzag D, Burger PC, Allen JC, Eberhart CG, Bar EE. BRAF alterations in primary glial and glioneuronal neoplasms of the central nervous system with identification of 2 novel KIAA1549:BRAF fusion variants. J Neuropathol Exp Neurol. 2012 Jan;71(1):66–72. doi: 10.1097/NEN.0b013e31823f2cb0. PMID: 22157620; PMCID: PMC4629834.

19. Sugiura Y, Nagaishi M. Clinical relevance of BRAF status in glial and glioneuronal tumors: A systematic review. J Clin Neurosci. 2019 Aug;66:196–201. doi: 10.1016/j.jocn.2019.05.014. Epub 2019 May 27. PMID: 31147232.

20. Di Nunno V, Gatto L, Tosoni A, Bartolini S, Franceschi E. Implications of BRAF V600E mutation in gliomas: Molecular considerations, prognostic value and treatment evolution. Front Oncol. 2023 Jan 4;12:1067252. doi: 10.3389/fonc.2022.1067252. PMID: 36686797; PMCID: PMC9846085.

21. Nguyen AT, Colin C, Nanni-Metellus I, Padovani L, Maurage CA, Varlet P, Miquel C, Uro-Coste E, Godfraind C, Lechapt-Zalcman E, Labrousse F, Gauchotte G, Silva K, Jouvet A, Figarella-Branger D; French GENOP Network. Evidence for BRAF V600E and H3F3A K27M double mutations in paediatric glial and glioneuronal tumours. Neuropathol Appl Neurobiol. 2015 Apr;41(3):403–8. doi: 10.1111/nan.12196. PMID: 25389051.

22. Trinder SM, McKay C, Power P, Topp M, Chan B, Valvi S, McCowage G, Govender D, Kirby M, Ziegler DS, Manoharan N, Hassall T, Kellie S, Heath J, Alvaro F, Wood P, Laughton S, Tsui K, Dodgshun A, Eisenstat DD, Endersby R, Luen SJ, Koh ES, Sim HW, Kong B, Gottardo NG, Whittle JR, Khuong-Quang DA, Hansford JR. BRAF-mediated brain tumors in adults and children: A review and the Australian and New Zealand experience. Front Oncol. 2023 Apr 14;13:1154246. doi: 10.3389/fonc.2023.1154246. PMID: 37124503; PMCID: PMC10140567.

23. Dimitriadis E, Alexiou GA, Tsotsou P, Simeonidi E, Stefanaki K, Patereli A, Prodromou N, Pandis N. BRAF alterations in pediatric low grade gliomas and mixed neuronal-glial tumors. J Neurooncol. 2013 Jul;113(3):353–8. doi: 10.1007/s11060-013-1131-5. Epub 2013 Apr 24. PMID: 23612919.

24. Hegedus B, Dasgupta B, Shin JE, Emnett RJ, Hart-Mahon EK, Elghazi L, Bernal-Mizrachi E, Gutmann DH. Neurofibromatosis-1 regulates neuronal and glial cell differentiation from neuroglial progenitors in vivo by both cAMP- and Ras-dependent mechanisms. Cell Stem Cell. 2007 Oct 11;1(4):443–57. doi: 10.1016/j.stem.2007.07.008. PMID: 18371380.

25. Helfferich J, Nijmeijer R, Brouwer OF, Boon M, Fock A, Hoving EW, Meijer L, den Dunnen WF, de Bont ES. Neurofibromatosis type 1 associated low grade gliomas: A comparison with sporadic low grade gliomas. Crit Rev Oncol Hematol. 2016 Aug;104:30–41. doi: 10.1016/j.critrevonc.2016.05.008. Epub 2016 May 21. PMID: 27263935.

26. Albers AC, Gutmann DH. Gliomas in patients with neurofibromatosis type 1. Expert Rev Neurother. 2009 Apr;9(4):535–9. doi: 10.1586/ern.09.4. PMID: 19344304.

27. Lee DY, Yeh TH, Emnett RJ, White CR, Gutmann DH. Neurofibromatosis-1 regulates neuroglial progenitor proliferation and glial differentiation in a brain region-specific manner. Genes Dev. 2010 Oct 15;24(20):2317–29. doi: 10.1101/gad.1957110. Epub 2010 Sep 28. PMID: 20876733; PMCID: PMC2956210.

28. D’Angelo F, Ceccarelli M, Tala, Garofano L, Zhang J, Frattini V, Caruso FP, Lewis G, Alfaro KD, Bauchet L, Berzero G, Cachia D, Cangiano M, Capelle L, de Groot J, DiMeco F, Ducray F, Farah W, Finocchiaro G, Goutagny S, Kamiya-Matsuoka C, Lavarino C, Loiseau H, Lorgis V, Marras CE, McCutcheon I, Nam DH, Ronchi S, Saletti V, Seizeur R, Slopis J, Suñol M, Vandenbos F, Varlet P, Vidaud D, Watts C, Tabar V, Reuss DE, Kim SK, Meyronet D, Mokhtari K, Salvador H, Bhat KP, Eoli M, Sanson M, Lasorella A, Iavarone A. The molecular landscape of glioma in patients with Neurofibromatosis 1. Nat Med. 2019 Jan;25(1):176–187. doi: 10.1038/s41591-018-0263-8. Epub 2018 Dec 10. PMID: 30531922; PMCID: PMC6857804.

29. Bajenaru ML, Zhu Y, Hedrick NM, Donahoe J, Parada LF, Gutmann DH. Astrocyte-specific inactivation of the neurofibromatosis 1 gene (NF1) is insufficient for astrocytoma formation. Mol Cell Biol. 2002 Jul;22(14):5100–13. doi: 10.1128/MCB.22.14.5100-5113.2002. PMID: 12077339; PMCID: PMC139771.

30. Gessi M, Moneim YA, Hammes J, Goschzik T, Scholz M, Denkhaus D, Waha A, Pietsch T. FGFR1 mutations in Rosette-forming glioneuronal tumors of the fourth ventricle. J Neuropathol Exp Neurol. 2014 Jun;73(6):580–4. doi: 10.1097/NEN.0000000000000080. PMID: 24806303.

31. Picca A, Berzero G, Bielle F, Touat M, Savatovsky J, Polivka M, Trisolini E, Meunier S, Schmitt Y, Idbaih A, Hoang-Xuan K, Delattre JY, Mokhtari K, Di Stefano AL, Sanson M. *FGFR1* actionable mutations, molecular specificities, and outcome of adult midline gliomas. Neurology. 2018 Jun 5;90(23):e2086–e2094. doi: 10.1212/WNL.0000000000005658. Epub 2018 May 4. PMID: 29728520.

32. Irschick R, Trost T, Karp G, Hausott B, Auer M, Claus P, Klimaschewski L. Sorting of the FGF receptor 1 in a human glioma cell line. Histochem Cell Biol. 2013 Jan;139(1):135–48. doi: 10.1007/s00418-012-1009-1. Epub 2012 Aug 18. PMID: 22903848.

33. Alshahrany N, Begum A, Siebzehnrubl D, Jimenez-Pascual A, Siebzehnrubl FA. Spatial distribution and functional relevance of FGFR1 and FGFR2 expression for glioblastoma tumor invasion. Cancer Lett. 2023 Sep 1;571:216349. doi: 10.1016/j.canlet.2023.216349. Epub 2023 Aug 12. PMID: 37579831; PMCID: PMC10840508.

34. Ma DK, Ponnusamy K, Song MR, Ming GL, Song H. Molecular genetic analysis of FGFR1 signalling reveals distinct roles of MAPK and PLCgamma1 activation for self-renewal of adult neural stem cells. Mol Brain. 2009 Jun 8;2:16. doi: 10.1186/1756-6606-2-16. PMID: 19505325; PMCID: PMC2700800.

35. Zhao M, Li D, Shimazu K, Zhou YX, Lu B, Deng CX. Fibroblast growth factor receptor-1 is required for long-term potentiation, memory consolidation, and neurogenesis. Biol Psychiatry. 2007 Sep 1;62(5):381–90. doi: 10.1016/j.biopsych.2006.10.019. Epub 2007 Jan 18. PMID: 17239352.

36. Smith JS, Tachibana I, Passe SM, Huntley BK, Borell TJ, Iturria N, O’Fallon JR, Schaefer PL, Scheithauer BW, James CD, Buckner JC, Jenkins RB. PTEN mutation, EGFR amplification, and outcome in patients with anaplastic astrocytoma and glioblastoma multiforme. J Natl Cancer Inst. 2001 Aug 15;93(16):1246–56. doi: 10.1093/jnci/93.16.1246. PMID: 11504770.

37. Knobbe CB, Merlo A, Reifenberger G. Pten signaling in gliomas. Neuro Oncol. 2002 Jul;4(3):196–211. PMID: 12084351; PMCID: PMC1920635.

38. Woods C, Flockton AR, Belkind-Gerson J. Phosphatase and Tensin Homolog Inhibition in Proteolipid Protein 1-Expressing Cells Stimulates Neurogenesis and Gliogenesis in the Postnatal Enteric Nervous System. Biomolecules. 2024 Mar 13;14(3):346. doi: 10.3390/biom14030346. PMID: 38540765; PMCID: PMC10967813.

39. Abe T, Terada K, Wakimoto H, Inoue R, Tyminski E, Bookstein R, Basilion JP, Chiocca EA. PTEN decreases in vivo vascularization of experimental gliomas in spite of proangiogenic stimuli. Cancer Res. 2003 May 1;63(9):2300–5. PMID: 12727853.

40. Giotta Lucifero A, Luzzi S. Immune Landscape in PTEN-Related Glioma Microenvironment: A Bioinformatic Analysis. Brain Sci. 2022 Apr 14;12(4):501. doi: 10.3390/brainsci12040501. PMID: 35448032; PMCID: PMC9029006.

41. Adachi J, Ohbayashi K, Suzuki T, Sasaki T. Cell cycle arrest and astrocytic differentiation resulting from PTEN expression in glioma cells. J Neurosurg. 1999 Nov;91(5):822–30. doi: 10.3171/jns.1999.91.5.0822. PMID: 10541240.

42. Jebelli JD, Hooper C, Garden GA, Pocock JM. Emerging roles of p53 in glial cell function in health and disease. Glia. 2012 Apr;60(4):515–25. doi: 10.1002/glia.22268. Epub 2011 Nov 21. PMID: 22105777; PMCID: PMC4195591.

43. Zhang Y, Dube C, Gibert M Jr, Cruickshanks N, Wang B, Coughlan M, Yang Y, Setiady I, Deveau C, Saoud K, Grello C, Oxford M, Yuan F, Abounader R. The p53 Pathway in Glioblastoma. Cancers (Basel). 2018 Sep 1;10(9):297. doi: 10.3390/cancers10090297. PMID: 30200436; PMCID: PMC6162501.

44. Mawrin C, Kirches E, Schneider-Stock R, Scherlach C, Vorwerk C, Von Deimling A, Van Landeghem F, Meyermann R, Bornemann A, Müller A, Romeike B, Stoltenburg-Didinger G, Wickboldt J, Pilz P, Dietzmann K. Analysis of TP53 and PTEN in gliomatosis cerebri. Acta Neuropathol. 2003 Jun;105(6):529–36. doi: 10.1007/s00401-003-0674-5. Epub 2003 Feb 26. PMID: 12734658.

45. Chung R, Whaley J, Kley N, Anderson K, Louis D, Menon A, Hettlich C, Freiman R, Hedley-Whyte ET, Martuza R, et al. TP53 gene mutations and 17p deletions in human astrocytomas. Genes Chromosomes Cancer. 1991 Sep;3(5):323–31. doi: 10.1002/gcc.2870030502. PMID: 1686725.

46. Faria MH, Neves Filho EH, Alves MK, Burbano RM, de Moraes Filho MO, Rabenhorst SH. TP53 mutations in astrocytic gliomas: an association with histological grade, TP53 codon 72 polymorphism and p53 expression. APMIS. 2012 Nov;120(11):882–9. doi: 10.1111/j.1600-0463.2012.02918.x. Epub 2012 May 18. PMID: 23009112.

47. Noor H, Briggs NE, McDonald KL, Holst J, Vittorio O. *TP53* Mutation Is a Prognostic Factor in Lower Grade Glioma and May Influence Chemotherapy Efficacy. Cancers (Basel). 2021 Oct 26;13(21):5362. doi: 10.3390/cancers13215362. PMID: 34771529; PMCID: PMC8582451.

48. Britsch S, Goerich DE, Riethmacher D, Peirano RI, Rossner M, Nave KA, Birchmeier C, Wegner M. The transcription factor Sox10 is a key regulator of peripheral glial development. Genes Dev. 2001 Jan 1;15(1):66–78. doi: 10.1101/gad.186601. PMID: 11156606; PMCID: PMC312607.

49. Kuhlbrodt K, Herbarth B, Sock E, Hermans-Borgmeyer I, Wegner M. Sox10, a novel transcriptional modulator in glial cells. J Neurosci. 1998 Jan 1;18(1):237–50. doi: 10.1523/JNEUROSCI.18-01-00237.1998. PMID: 9412504; PMCID: PMC6793382.

50. Kuhlbrodt K, Herbarth B, Sock E, Hermans-Borgmeyer I, Wegner M. Sox10, a novel transcriptional modulator in glial cells. J Neurosci. 1998 Jan 1;18(1):237–50. doi: 10.1523/JNEUROSCI.18-01-00237.1998. PMID: 9412504; PMCID: PMC6793382.

51. Paratore C, Goerich DE, Suter U, Wegner M, Sommer L. Survival and glial fate acquisition of neural crest cells are regulated by an interplay between the transcription factor Sox10 and extrinsic combinatorial signaling. Development. 2001 Oct;128(20):3949–61. doi: 10.1242/dev.128.20.3949. PMID: 11641219.

52. Kleinschmidt-DeMasters BK, Donson AM, Richmond AM, Pekmezci M, Tihan T, Foreman NK. SOX10 Distinguishes Pilocytic and Pilomyxoid Astrocytomas From Ependymomas but Shows No Differences in Expression Level in Ependymomas From Infants Versus Older Children or Among Molecular Subgroups. J Neuropathol Exp Neurol. 2016 Apr;75(4):295–8. doi: 10.1093/jnen/nlw010. Epub 2016 Mar 4. PMID: 26945037; PMCID: PMC5009481.

53. Kordes U, Hagel C. Expression of SOX9 and SOX10 in central neuroepithelial tumor. J Neurooncol. 2006 Nov;80(2):151–5. doi: 10.1007/s11060-006-9180-7. Epub 2006 Jun 22. PMID: 16791471.

54. Chen KS, Bridges CR, Lynton Z, Lim JWC, Stringer BW, Rajagopal R, Wong KT, Ganesan D, Ariffin H, Day BW, Richards LJ, Bunt J. Transcription factors NFIA and NFIB induce cellular differentiation in high-grade astrocytoma. J Neurooncol. 2020 Jan;146(1):41–53. doi: 10.1007/s11060-019-03352-3. Epub 2019 Nov 23. PMID: 31760595.

55. Cimino PJ, Ketchum C, Turakulov R, Singh O, Abdullaev Z, Giannini C, Pytel P, Lopez GY, Colman H, Nasrallah MP, Santi M, Fernandes IL, Nirschl J, Dahiya S, Neill S, Solomon D, Perez E, Capper D, Mani H, Caccamo D, Ball M, Badruddoja M, Chkheidze R, Camelo-Piragua S, Fullmer J, Alexandrescu S, Yeaney G, Eberhart C, Martinez-Lage M, Chen J, Zach L, Kleinschmidt-DeMasters BK, Hefti M, Lopes MB, Nuechterlein N, Horbinski C, Rodriguez FJ, Quezado M, Pratt D, Aldape K. Expanded analysis of high-grade astrocytoma with piloid features identifies an epigenetically and clinically distinct subtype associated with neurofibromatosis type 1. Acta Neuropathol. 2023 Jan;145(1):71–82. doi: 10.1007/s00401-022-02513-5. Epub 2022 Oct 22. PMID: 36271929; PMCID: PMC9844520.

56. Song HR, Gonzalez-Gomez I, Suh GS, Commins DL, Sposto R, Gilles FH, Deneen B, Erdreich-Epstein A. Nuclear factor IA is expressed in astrocytomas and is associated with improved survival. Neuro Oncol. 2010 Feb;12(2):122–32. doi: 10.1093/neuonc/nop044. Epub 2010 Jan 25. PMID: 20150379; PMCID: PMC2940580.

57. Deneen B, Ho R, Lukaszewicz A, Hochstim CJ, Gronostajski RM, Anderson DJ. The transcription factor NFIA controls the onset of gliogenesis in the developing spinal cord. Neuron. 2006 Dec 21;52(6):953–68. doi: 10.1016/j.neuron.2006.11.019. PMID: 17178400.

58. Tchieu J, Calder EL, Guttikonda SR, Gutzwiller EM, Aromolaran KA, Steinbeck JA, Goldstein PA, Studer L. NFIA is a gliogenic switch enabling rapid derivation of functional human astrocytes from pluripotent stem cells. Nat Biotechnol. 2019 Mar;37(3):267–275. doi: 10.1038/s41587-019-0035-0. Epub 2019 Feb 25. PMID: 30804533; PMCID: PMC6591152.

59. Lahti L, Volakakis N, Gillberg L, Yaghmaeian Salmani B, Tiklová K, Kee N, Lundén-Miguel H, Werkman M, Piper M, Gronostajski R, Perlmann T. Sox9 and Nfi transcription factors regulate the timing of neurogenesis and ependymal maturation in dopamine progenitors. Development. 2025 Feb 25:dev.204421. doi: 10.1242/dev.204421. Epub ahead of print. PMID: 39995267.

60. Mizoguchi M, Betensky RA, Batchelor TT, Bernay DC, Louis DN, Nutt CL. Activation of STAT3, MAPK, and AKT in malignant astrocytic gliomas: correlation with EGFR status, tumor grade, and survival. J Neuropathol Exp Neurol. 2006 Dec;65(12):1181–8. doi: 10.1097/01.jnen.0000248549.14962.b2. PMID: 17146292.

61. Liang Q, Ma C, Zhao Y, Gao G, Ma J. Inhibition of STAT3 reduces astrocytoma cell invasion and constitutive activation of STAT3 predicts poor prognosis in human astrocytoma. PLoS One. 2013 Dec 30;8(12):e84723. doi: 10.1371/journal.pone.0084723. PMID: 24386409; PMCID: PMC3875539.

62. Fu W, Hou X, Dong L, Hou W. Roles of STAT3 in the pathogenesis and treatment of glioblastoma. Front Cell Dev Biol. 2023 Feb 27;11:1098482. doi: 10.3389/fcell.2023.1098482. PMID: 36923251; PMCID: PMC10009693.

63. de la Iglesia N, Puram SV, Bonni A. STAT3 regulation of glioblastoma pathogenesis. Curr Mol Med. 2009 Jun;9(5):580–90. doi: 10.2174/156652409788488739. PMID: 19601808; PMCID: PMC2712135.

64. Hagemann TL, Coyne S, Levin A, Wang L, Feany MB, Messing A. STAT3 Drives GFAP Accumulation and Astrocyte Pathology in a Mouse Model of Alexander Disease. Cells. 2023 Mar 23;12(7):978. doi: 10.3390/cells12070978. PMID: 37048051; PMCID: PMC10093589.

65. Acarin L, González B, Castellano B. STAT3 and NFkappaB activation precedes glial reactivity in the excitotoxically injured young cortex but not in the corresponding distal thalamic nuclei. J Neuropathol Exp Neurol. 2000 Feb;59(2):151–63. doi: 10.1093/jnen/59.2.151. PMID: 10749104.

66. Zhou YH, Tan F, Hess KR, Yung WK. The expression of PAX6, PTEN, vascular endothelial growth factor, and epidermal growth factor receptor in gliomas: relationship to tumor grade and survival. Clin Cancer Res. 2003 Aug 15;9(9):3369–75. PMID: 12960124.

67. Sansom SN, Griffiths DS, Faedo A, Kleinjan DJ, Ruan Y, Smith J, van Heyningen V, Rubenstein JL, Livesey FJ. The level of the transcription factor Pax6 is essential for controlling the balance between neural stem cell self-renewal and neurogenesis. PLoS Genet. 2009 Jun;5(6):e1000511. doi: 10.1371/journal.pgen.1000511. Epub 2009 Jun 12. PMID: 19521500; PMCID: PMC2686252.

68. Kallur T, Gisler R, Lindvall O, Kokaia Z. Pax6 promotes neurogenesis in human neural stem cells. Mol Cell Neurosci. 2008 Aug;38(4):616–28. doi: 10.1016/j.mcn.2008.05.010. Epub 2008 May 22. PMID: 18595732.

69. Shohayeb B, Cooper HM. The ups and downs of Pax6 in neural stem cells. J Biol Chem. 2023 May;299(5):104680. doi: 10.1016/j.jbc.2023.104680. Epub 2023 Apr 5. PMID: 37028762; PMCID: PMC10164895.

70. Curto GG, Nieto-Estévez V, Hurtado-Chong A, Valero J, Gómez C, Alonso JR, Weruaga E, Vicario-Abejón C. Pax6 is essential for the maintenance and multi-lineage differentiation of neural stem cells, and for neuronal incorporation into the adult olfactory bulb. Stem Cells Dev. 2014 Dec 1;23(23):2813–30. doi: 10.1089/scd.2014.0058. Epub 2014 Sep 17. PMID: 25117830; PMCID: PMC4235597.

71. Zhou YH, Wu X, Tan F, Shi YX, Glass T, Liu TJ, Wathen K, Hess KR, Gumin J, Lang F, Yung WK. PAX6 suppresses growth of human glioblastoma cells. J Neurooncol. 2005 Feb;71(3):223–9. doi: 10.1007/s11060-004-1720-4. PMID: 15735909.

72. Byrns CN, Saikumar J, Bonini NM. Glial AP1 is activated with aging and accelerated by traumatic brain injury. Nat Aging. 2021 Jul;1(7):585–597. doi: 10.1038/s43587-021-00072-0. Epub 2021 Jul 8. PMID: 34723199; PMCID: PMC8553014.

73. Brenner M, Messing A, Olsen ML. AP-1 and the injury response of the GFAP gene. J Neurosci Res. 2019 Feb;97(2):149–161. doi: 10.1002/jnr.24338. Epub 2018 Oct 22. PMID: 30345544; PMCID: PMC6289842.

74. Assimakopoulou M, Varakis J. AP-1 and heat shock protein 27 expression in human astrocytomas. J Cancer Res Clin Oncol. 2001 Dec;127(12):727–32. doi: 10.1007/s004320100280. PMID: 11768612.

75. Bhardwaj R, Suzuki A, Leland P, Joshi BH, Puri RK. Identification of a novel role of IL-13Rα2 in human Glioblastoma multiforme: interleukin-13 mediates signal transduction through AP-1 pathway. J Transl Med. 2018 Dec 20;16(1):369. doi: 10.1186/s12967-018-1746-6. PMID: 30572904; PMCID: PMC6302477.

76. Li ZH, Guan YL, Liu Q, Wang Y, Cui R, Wang YJ. Astrocytoma progression scoring system based on the WHO 2016 criteria. Sci Rep. 2019 Jan 14;9(1):96. doi: 10.1038/s41598-018-36471-4. PMID: 30643174; PMCID: PMC6331604.

77. Jeyapalan JN, Doctor GT, Jones TA, Alberman SN, Tep A, Haria CM, Schwalbe EC, Morley IC, Hill AA, LeCain M, Ottaviani D, Clifford SC, Qaddoumi I, Tatevossian RG, Ellison DW, Sheer D. DNA methylation analysis of paediatric low-grade astrocytomas identifies a tumour-specific hypomethylation signature in pilocytic astrocytomas. Acta Neuropathol Commun. 2016 May 27;4(1):54. doi: 10.1186/s40478-016-0323-6. PMID: 27229157; PMCID: PMC4882864.

78. Ono K, Takebayashi H, Ikenaka K. Olig2 transcription factor in the developing and injured forebrain; cell lineage and glial development. Mol Cells. 2009 Apr 30;27(4):397–401. doi: 10.1007/s10059-009-0067-2. Epub 2009 Apr 13. PMID: 19390819.

79. Cai J, Chen Y, Cai WH, Hurlock EC, Wu H, Kernie SG, Parada LF, Lu QR. A crucial role for Olig2 in white matter astrocyte development. Development. 2007 May;134(10):1887–99. doi: 10.1242/dev.02847. Epub 2007 Apr 11. PMID: 17428828.

80. Ligon KL, Alberta JA, Kho AT, Weiss J, Kwaan MR, Nutt CL, Louis DN, Stiles CD, Rowitch DH. The oligodendroglial lineage marker OLIG2 is universally expressed in diffuse gliomas. J Neuropathol Exp Neurol. 2004 May;63(5):499–509. doi: 10.1093/jnen/63.5.499. PMID: 15198128.

81. Otero JJ, Rowitch D, Vandenberg S. OLIG2 is differentially expressed in pediatric astrocytic and in ependymal neoplasms. J Neurooncol. 2011 Sep;104(2):423–38. doi: 10.1007/s11060-010-0509-x. Epub 2010 Dec 31. PMID: 21193945; PMCID: PMC3161192.

82. Ishizawa K, Komori T, Shimada S, Hirose T. Olig2 and CD99 are useful negative markers for the diagnosis of brain tumors. Clin Neuropathol. 2008 May-Jun;27(3):118–28. doi: 10.5414/npp27118. PMID: 18552083.

83. Bouchart C, Trépant AL, Hein M, Van Gestel D, Demetter P. Prognostic impact of glioblastoma stem cell markers OLIG2 and CCND2. Cancer Med. 2020 Feb;9(3):1069–1078. doi: 10.1002/cam4.2592. Epub 2019 Sep 30. PMID: 31568682; PMCID: PMC6997071.

84. Iroegbu JD, Ijomone OK, Femi-Akinlosotu OM, Ijomone OM. ERK/MAPK signalling in the developing brain: Perturbations and consequences. Neurosci Biobehav Rev. 2021 Dec;131:792–805. doi: 10.1016/j.neubiorev.2021.10.009. Epub 2021 Oct 8. PMID: 34634357.

85. Baumann NS, Sears JC, Broadie K. Experience-dependent MAPK/ERK signaling in glia regulates critical period remodeling of synaptic glomeruli. Cell Signal. 2024 Aug;120:111224. doi: 10.1016/j.cellsig.2024.111224. Epub 2024 May 11. PMID: 38740233; PMCID: PMC11459659.

86. Albert-Gascó H, Ros-Bernal F, Castillo-Gómez E, Olucha-Bordonau FE. MAP/ERK Signaling in Developing Cognitive and Emotional Function and Its Effect on Pathological and Neurodegenerative Processes. Int J Mol Sci. 2020 Jun 23;21(12):4471. doi: 10.3390/ijms21124471. PMID: 32586047; PMCID: PMC7352860.

87. Salles D, Santino SF, Ribeiro DA, Malinverni ACM, Stávale JN. The involvement of the MAPK pathway in pilocytic astrocytomas. Pathol Res Pract. 2022 Apr;232:153821. doi: 10.1016/j.prp.2022.153821. Epub 2022 Feb 25. PMID: 35231859.

88. Forshew T, Tatevossian RG, Lawson AR, Ma J, Neale G, Ogunkolade BW, Jones TA, Aarum J, Dalton J, Bailey S, Chaplin T, Carter RL, Gajjar A, Broniscer A, Young BD, Ellison DW, Sheer D. Activation of the ERK/MAPK pathway: a signature genetic defect in posterior fossa pilocytic astrocytomas. J Pathol. 2009 Jun;218(2):172–81. doi: 10.1002/path.2558. PMID: 19373855.

89. Jones DT, Gronych J, Lichter P, Witt O, Pfister SM. MAPK pathway activation in pilocytic astrocytoma. Cell Mol Life Sci. 2012 Jun;69(11):1799–811. doi: 10.1007/s00018-011-0898-9. Epub 2011 Dec 13. PMID: 22159586; PMCID: PMC3350769.

90. Ma Q, Chen G, Li Y, Guo Z, Zhang X. The molecular genetics of PI3K/PTEN/AKT/mTOR pathway in the malformations of cortical development. Genes Dis. 2023 Jul 16;11(5):101021. doi: 10.1016/j.gendis.2023.04.041. PMID: 39006182; PMCID: PMC11245990.

91. Glaviano A, Foo ASC, Lam HY, Yap KCH, Jacot W, Jones RH, Eng H, Nair MG, Makvandi P, Geoerger B, Kulke MH, Baird RD, Prabhu JS, Carbone D, Pecoraro C, Teh DBL, Sethi G, Cavalieri V, Lin KH, Javidi-Sharifi NR, Toska E, Davids MS, Brown JR, Diana P, Stebbing J, Fruman DA, Kumar AP. PI3K/AKT/mTOR signaling transduction pathway and targeted therapies in cancer. Mol Cancer. 2023 Aug 18;22(1):138. doi: 10.1186/s12943-023-01827-6. PMID: 37596643; PMCID: PMC10436543.

92. Wu YT, Tan HL, Huang Q, Ong CN, Shen HM. Activation of the PI3K-Akt-mTOR signaling pathway promotes necrotic cell death via suppression of autophagy. Autophagy. 2009 Aug;5(6):824–34. doi: 10.4161/auto.9099. Epub 2009 Aug 26. PMID: 19556857.

93. Porta C, Paglino C, Mosca A. Targeting PI3K/Akt/mTOR Signaling in Cancer. Front Oncol. 2014 Apr 14;4:64. doi: 10.3389/fonc.2014.00064. PMID: 24782981; PMCID: PMC3995050.

94. Omolekan TO, Chamcheu JC, Buerger C, Huang S. PI3K/AKT/mTOR Signaling Network in Human Health and Diseases. Cells. 2024 Sep 6;13(17):1500. doi: 10.3390/cells13171500. PMID: 39273070; PMCID: PMC11394329.

95. Barzegar Behrooz A, Talaie Z, Jusheghani F, Łos MJ, Klonisch T, Ghavami S. Wnt and PI3K/Akt/mTOR Survival Pathways as Therapeutic Targets in Glioblastoma. Int J Mol Sci. 2022 Jan 25;23(3):1353. doi: 10.3390/ijms23031353. PMID: 35163279; PMCID: PMC8836096.

96. Justicia C, Gabriel C, Planas AM. Activation of the JAK/STAT pathway following transient focal cerebral ischemia: signaling through Jak1 and Stat3 in astrocytes. Glia. 2000 May;30(3):253–70. doi: 10.1002/(sici)1098-1136(200005)30:3<253::aid-glia5>3.0.co;2-o. PMID: 10756075.

97. Na YJ, Jin JK, Kim JI, Choi EK, Carp RI, Kim YS. JAK-STAT signaling pathway mediates astrogliosis in brains of scrapie-infected mice. J Neurochem. 2007 Oct;103(2):637–49. doi: 10.1111/j.1471-4159.2007.04769.x. PMID: 17897356.

98. Ceyzériat K, Abjean L, Carrillo-de Sauvage MA, Ben Haim L, Escartin C. The complex STATes of astrocyte reactivity: How are they controlled by the JAK-STAT3 pathway? Neuroscience. 2016 Aug 25;330:205–18. doi: 10.1016/j.neuroscience.2016.05.043. Epub 2016 May 27. PMID: 27241943.

99. Rusek M, Smith J, El-Khatib K, Aikins K, Czuczwar SJ, Pluta R. The Role of the JAK/STAT Signaling Pathway in the Pathogenesis of Alzheimer’s Disease: New Potential Treatment Target. Int J Mol Sci. 2023 Jan 3;24(1):864. doi: 10.3390/ijms24010864. PMID: 36614305; PMCID: PMC9821184.

100. Ben Haim L, Ceyzériat K, Carrillo-de Sauvage MA, Aubry F, Auregan G, Guillermier M, Ruiz M, Petit F, Houitte D, Faivre E, Vandesquille M, Aron-Badin R, Dhenain M, Déglon N, Hantraye P, Brouillet E, Bonvento G, Escartin C. The JAK/STAT3 pathway is a common inducer of astrocyte reactivity in Alzheimer’s and Huntington’s diseases. J Neurosci. 2015 Feb 11;35(6):2817–29. doi: 10.1523/JNEUROSCI.3516-14.2015. PMID: 25673868; PMCID: PMC6605603.

101. Nicolas CS, Amici M, Bortolotto ZA, Doherty A, Csaba Z, Fafouri A, Dournaud P, Gressens P, Collingridge GL, Peineau S. The role of JAK-STAT signaling within the CNS. JAKSTAT. 2013 Jan 1;2(1):e22925. doi: 10.4161/jkst.22925. PMID: 24058789; PMCID: PMC3670265.

102. Hill SA, Fu M, Garcia ADR. Sonic hedgehog signaling in astrocytes. Cell Mol Life Sci. 2021 Feb;78(4):1393–1403. doi: 10.1007/s00018-020-03668-8. Epub 2020 Oct 20. PMID: 33079226; PMCID: PMC7904711.

103. Garcia ADR. New Tricks for an Old (Hedge)Hog: Sonic Hedgehog Regulation of Astrocyte Function. Cells. 2021 May 30;10(6):1353. doi: 10.3390/cells10061353. PMID: 34070740; PMCID: PMC8228508.

104. Michinaga S, Hishinuma S, Koyama Y. Roles of astrocytic sonic hedgehog production and its signal for regulation of the blood-brain barrier permeability. Vitam Horm. 2024;126:97–111. doi: 10.1016/bs.vh.2024.04.006. Epub 2024 May 19. PMID: 39029978.

105. Xie Y, Kuan AT, Wang W, Herbert ZT, Mosto O, Olukoya O, Adam M, Vu S, Kim M, Tran D, Gómez N, Charpentier C, Sorour I, Lacey TE, Tolstorukov MY, Sabatini BL, Lee WA, Harwell CC. Astrocyte-neuron crosstalk through Hedgehog signaling mediates cortical synapse development. Cell Rep. 2022 Feb 22;38(8):110416. doi: 10.1016/j.celrep.2022.110416. PMID: 35196485; PMCID: PMC8962654.

106. Gingrich EC, Case K, Garcia ADR. A subpopulation of astrocyte progenitors defined by Sonic hedgehog signaling. Neural Dev. 2022 Jan 14;17(1):2. doi: 10.1186/s13064-021-00158-w. PMID: 35027088; PMCID: PMC8759290.

107. Wireko AA, Ben-Jaafar A, Kong JSH, Mannan KM, Sanker V, Rosenke SL, Boye ANA, Nkrumah-Boateng PA, Poornaselvan J, Shah MH, Abdul-Rahman T, Atallah O. Sonic hedgehog signalling pathway in CNS tumours: its role and therapeutic implications. Mol Brain. 2024 Nov 20;17(1):83. doi: 10.1186/s13041-024-01155-w. PMID: 39568072; PMCID: PMC11580395.

108. Ge W, Martinowich K, Wu X, He F, Miyamoto A, Fan G, Weinmaster G, Sun YE. Notch signaling promotes astrogliogenesis via direct CSL-mediated glial gene activation. J Neurosci Res. 2002 Sep 15;69(6):848–60. doi: 10.1002/jnr.10364. PMID: 12205678.

109. Givogri MI, de Planell M, Galbiati F, Superchi D, Gritti A, Vescovi A, de Vellis J, Bongarzone ER. Notch signaling in astrocytes and neuroblasts of the adult subventricular zone in health and after cortical injury. Dev Neurosci. 2006;28(1-2):81–91. doi: 10.1159/000090755. PMID: 16508306.

110. Zhang Y, He K, Wang F, Li X, Liu D. Notch-1 signaling regulates astrocytic proliferation and activation after hypoxia exposure. Neurosci Lett. 2015 Aug 31;603:12–8. doi: 10.1016/j.neulet.2015.07.009. Epub 2015 Jul 13. PMID: 26182882.

111. Wilhelmsson U, Faiz M, de Pablo Y, Sjöqvist M, Andersson D, Widestrand A, Potokar M, Stenovec M, Smith PL, Shinjyo N, Pekny T, Zorec R, Ståhlberg A, Pekna M, Sahlgren C, Pekny M. Astrocytes negatively regulate neurogenesis through the Jagged1-mediated Notch pathway. Stem Cells. 2012 Oct;30(10):2320–9. doi: 10.1002/stem.1196. PMID: 22887872.

112. Brandt WD, Schreck KC, Bar EE, Taylor I, Marchionni L, Raabe E, Eberhart CG, Rodriguez FJ. Notch signaling activation in pediatric low-grade astrocytoma. J Neuropathol Exp Neurol. 2015 Feb;74(2):121–31. doi: 10.1097/NEN.0000000000000155. PMID: 25575134; PMCID: PMC4357229.

113. Xu P, Yu S, Jiang R, Kang C, Wang G, Jiang H, Pu P. Differential expression of Notch family members in astrocytomas and medulloblastomas. Pathol Oncol Res. 2009 Dec;15(4):703–10. doi: 10.1007/s12253-009-9173-x. Epub 2009 May 8. PMID: 19424825.

